# Longitudinal multi-omic signatures of ARDS and sepsis inflammatory phenotypes identify key pathways associated with mortality

**DOI:** 10.1101/2025.05.07.25327117

**Authors:** Narges Alipanah-Lechner, Lucile Neyton, Pratik Sinha, Carolyn Leroux, Kim Bardillon, Sidney A. Carrillo, Suzanna Chak, Olivia Chao, Taarini Hariharan, Carolyn Henrickson, Kirsten Kangelaris, Charles R. Langelier, Deanna Lee, Chelsea Lin, Kathleen Liu, Liam Magee, Angelika Ringor, Aartik Sarma, Emma Schmiege, Natasha Spottiswoode, Kathryn Sullivan, Melanie F. Weingart, Andrew Willmore, Hanjing Zhuo, Angela J. Rogers, Kathleen A. Stringer, Michael A. Matthay, Carolyn S. Calfee

## Abstract

Critically ill patients with acute respiratory distress syndrome (ARDS) and sepsis exhibit distinct inflammatory phenotypes with divergent clinical outcomes and apparent heterogeneity of treatment effects, but the underlying molecular mechanisms remain poorly understood. These phenotypes, derived from clinical data and protein biomarkers, were associated with metabolic differences in a prior pilot study. This study investigated the metabolomic and transcriptomic differences between Hyperinflammatory and Hypoinflammatory phenotypes through integrative multi-omics analysis of blood samples from ARDS patients in the ROSE trial. Multi-omics integration revealed three molecular signatures strongly associated with the Hyperinflammatory phenotype and with mortality: enhanced innate immune activation coupled with increased glycolysis, hepatic dysfunction and immune dysfunction paired with impaired fatty acid beta-oxidation, and interferon program suppression coupled with altered mitochondrial respiration. A fourth molecular signature, not associated with inflammatory phenotype, identified redox impairment and cell proliferation pathways associated with mortality. Integrated multi-omics analysis <u>within</u> each inflammatory phenotype revealed distinct pathways associated with mortality. All mortality-associated molecular signatures including those within phenotypes were validated in an independent cohort of critically ill patients with sepsis (EARLI). These findings reveal distinct molecular mechanisms underlying ARDS/sepsis phenotypes and suggest potential therapeutic targets for precise treatment strategies in critical illness.

## INTRODUCTION

The acute respiratory distress syndrome (ARDS) and sepsis are devastating critical illness syndromes with unacceptably high mortality rates approaching 40-50% in the United States (*1, 2*). A significant challenge to developing effective treatments has been the marked heterogeneity in clinical presentation, underlying biology, and treatment responses among affected patients (*3, 4*).

Recent advances in molecular phenotyping have identified reproducible subgroups of ARDS and sepsis patients with distinct pathobiology. Latent class analyses (LCA) of clinical and plasma protein data consistently reveal two predominant phenotypes: a “Hyperinflammatory” phenotype characterized by elevated plasma inflammatory protein biomarkers, shock, and higher mortality, and a “Hypoinflammatory” phenotype with relatively lower inflammatory protein biomarkers and better outcomes (*5–10*). These phenotypes, identified across multiple ARDS and sepsis cohorts, demonstrate differential therapeutic responses in secondary analyses of randomized trials, suggesting they represent endotypes with distinct disease mechanisms (*5, 11, 12*). Clinical trials incorporating prospective phenotyping are being developed, including the PANTHER trial, which will start enrolling in mid-2025 (*13*). However, the biological processes driving each phenotype and mechanisms underlying unfavorable outcomes within each phenotype remain poorly understood. While protein biomarker studies have provided valuable insights into inflammatory patterns, they capture only a small fraction of the complex molecular landscape. Previous metabolic profiling of 93 patients with ARDS demonstrated that the Hyperinflammatory phenotype exhibits reduced circulating lipids and a glycolytic shift, while transcriptomic analyses revealed increased expression of genes related to the innate immune response and tissue remodeling and reduced interferon signaling (*10, 14*). However, isolated - omic approaches may miss critical interactions between cellular programming and systemic metabolism essential for understanding disease processes and treatment responses.

In this study, we applied longitudinal multi-omics profiling to characterize the molecular basis of ARDS/sepsis inflammatory phenotypes and identify mechanisms associated with poor outcomes. We hypothesized that these phenotypes would demonstrate distinct metabolic profiles and that integrated metabolomic-transcriptomic analysis would reveal novel outcome-associated mechanisms with therapeutic potential. By simultaneously measuring the metabolome and transcriptome at two timepoints in a large ARDS cohort, we aimed to: (1) identify novel metabolic differences between inflammatory phenotypes, (2) characterize coordinated metabolomic-transcriptomic signatures contributing to heterogeneity, (3) determine temporal stability of these patterns, and (4) uncover potentially targetable pathways associated with mortality. This comprehensive molecular characterization aims to advance our understanding of ARDS/sepsis heterogeneity and identify novel therapeutic approaches tailored to specific patient subgroups.

## RESULTS

### LCA phenotypes have distinct metabolic profiles

We first asked whether ethylenediaminetetraacetic acid (EDTA) plasma metabolites would be different between latent class analysis (LCA)-defined ARDS phenotypes. We evaluated patients from the ROSE trial of neuromuscular blockade for the treatment of moderate-to-severe ARDS (**Fig. 1A**), who had previously undergone LCA phenotyping using plasma protein biomarkers and clinical variables (*10, 15*). We randomly selected 80 patients in each phenotype with high phenotype membership probability (>0.9) (**Fig. S1**). These 160 total patients had a median age of 58.5 (IQR 47 to 68), were predominantly male (64%), and racially identified as white (78%), with equal proportions randomized to neuromuscular blockade across phenotypes (**Table S1**). The Hyperinflammatory group exhibited lower median body mass index (BMI), higher APACHEIII scores, reduced glomerular filtration rate (GFR), and higher prevalence of comorbid liver disease and leukemia. Corticosteroid administration rates were identical between phenotypes (24%). Consistent with previous studies, Hyperinflammatory patients more frequently required vasopressors at enrollment (86% vs 21%), experienced more than twice the mortality at 28 and 90 days, and had significantly fewer ventilator-, ICU-, and hospital-free days. Pneumonia was the predominant ARDS etiology in both phenotypes, while all patients with extrapulmonary sepsis-induced ARDS belonged to the Hyperinflammatory group.

**Figure 1.**
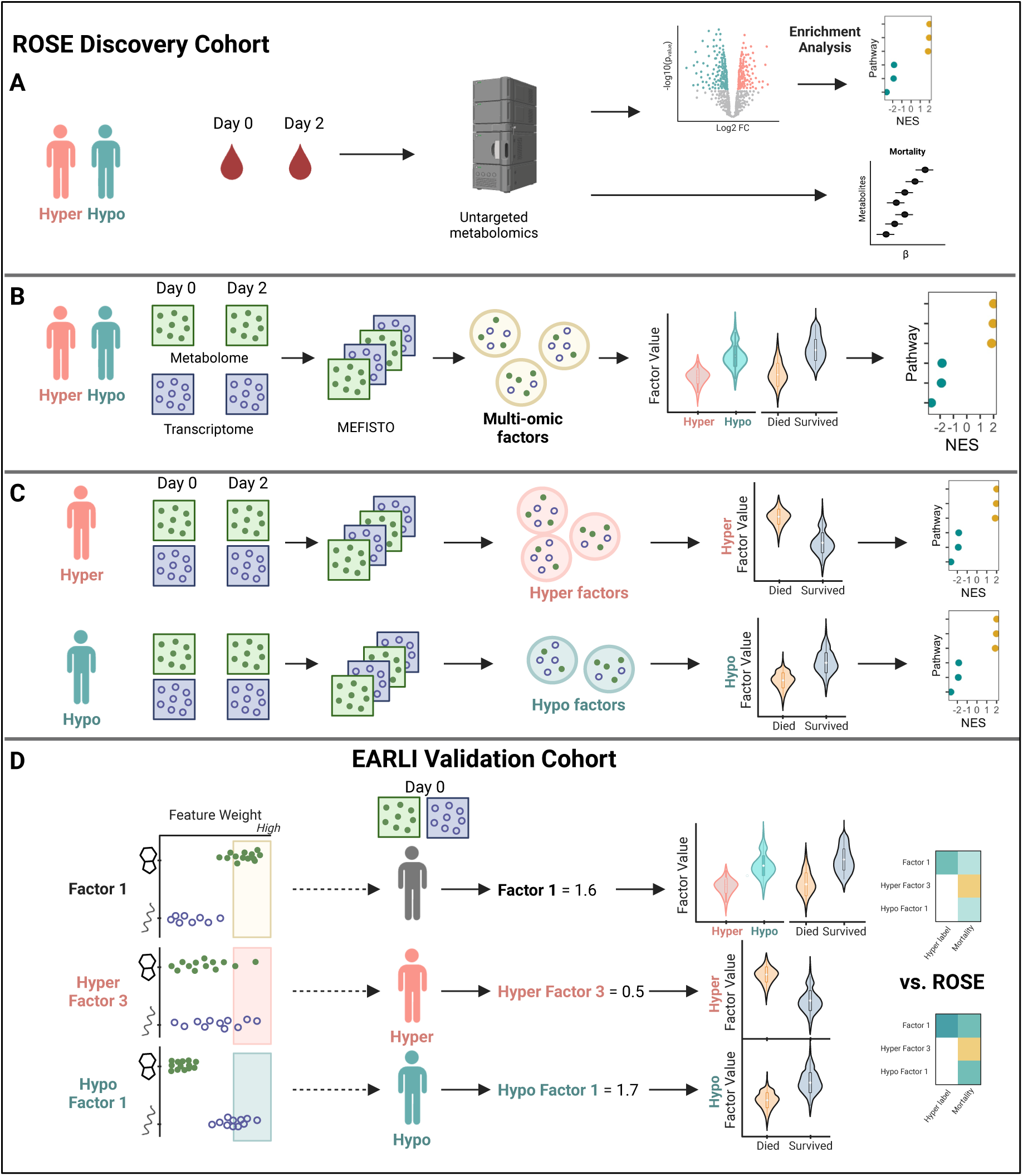
Study overview. (**A**) Day 0 and Day 2 EDTA plasma from ROSE study participants underwent untargeted metabolic profiling to determine differences between latent class analysis (LCA) based inflammatory phenotypes. (**B**) Longitudinal whole blood transcriptomic data and metabolomic data were analyzed using an unsupervised multi-modal factor analysis (MEFISTO) and the predominant sources of biological heterogeneity in the data of clinical relevance were assessed. (**C**) MEFISTO was applied separately to each phenotype to determine signatures related to mortality within each phenotype. (**D**) The highest weighted features (metabolite or gene) by absolute value within each multi-omic factor of interest were used to calculate factor weights for patients in an observational cohort study (EARLI). The association of factor weights with LCA phenotypes and outcomes in the validation cohort was assessed.

Untargeted metabolic profiling identified 1,378 known metabolites (**Fig. S2**). Differential abundance analysis using limma with adjustment for potential confounders identified 541 metabolites significantly different between phenotypes at Day 0, with substantial differences across all metabolic classes (**Fig. 2A-B**). Similar analysis at Day 2 revealed 496 significantly different metabolites, largely overlapping with Day 0 findings (**Fig. 2C**). Metabolite enrichment analysis highlighted 60 dysregulated pathways at Day 0 and 56 at Day 2, totaling 74 unique metabolic pathways (**Fig. 2D, Fig. S3**). The top 20 most differentially abundant metabolites belonged to lipids and amino acid classes, though the highest proportion of differentially abundant metabolites were related to energy production at both timepoints (**Table S2-S3**).

**Figure 2.**
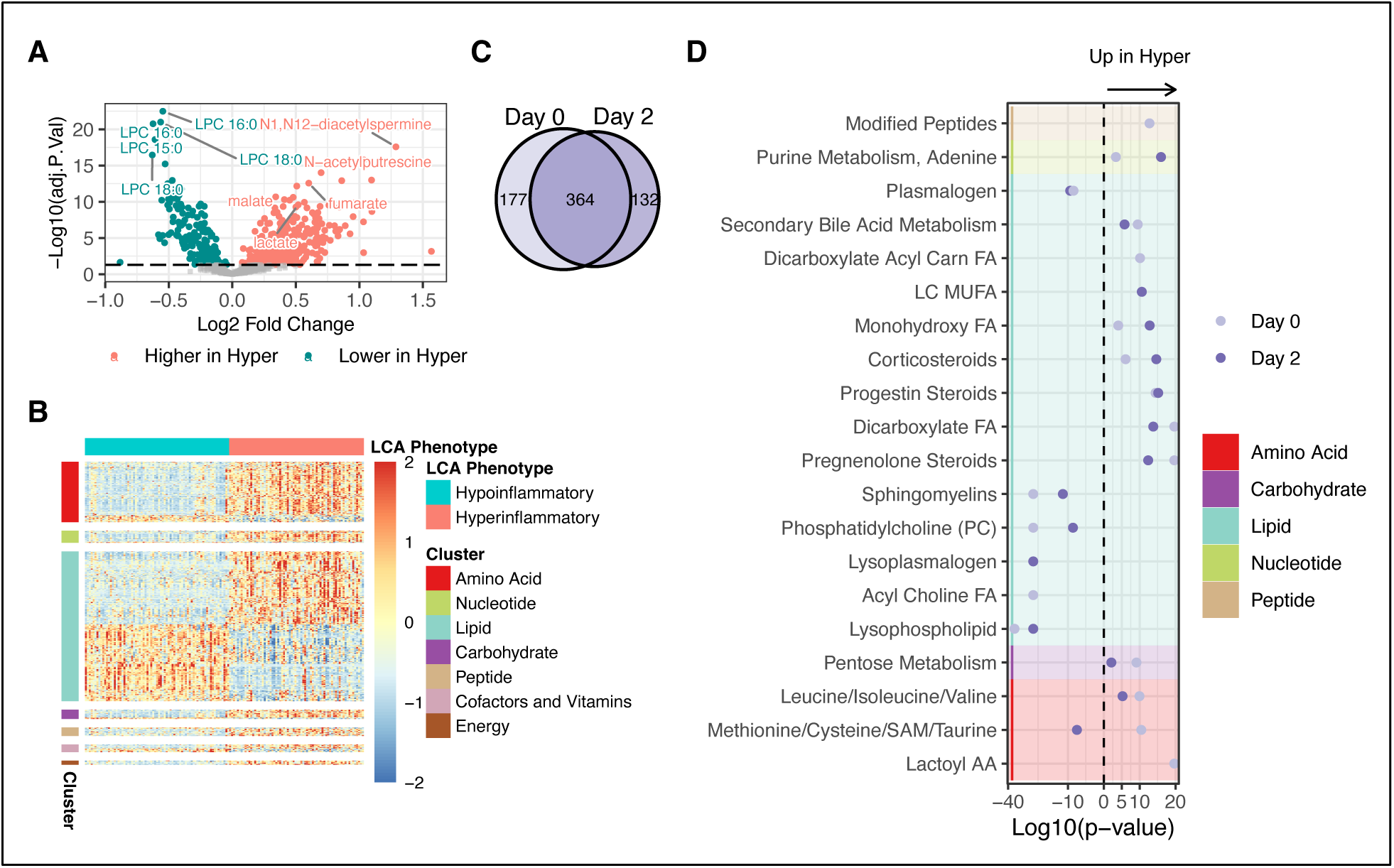
Metabolic profiling of latent class analysis (LCA) phenotypes. (**A**) Volcano plot showing differentially abundant metabolites between Hyperinflammatory and Hypoinflammatory ARDS at Day 0, determined by limma adjusted for covariates (age, sex, BMI, medications, liver disease, and GFR). (**B**) Heatmap of differentially abundant metabolites by LCA phenotype at Day 0 as determined by limma with adjustment for aforementioned covariates. Z-scaled log-transformed metabolite intensities are grouped by phenotype. (**C**) Venn diagram showing overlap of differentially abundant metabolites at Day 0 and Day 2 (Day 2 also adjusted for randomization arm). (**D**) Metabolite pathway enrichment analysis comparing Hyperinflammatory vs Hypoinflammatory groups at Day 0 and Day 2. X-axis shows signed log10(p-value), with positive values indicating positive enrichment in Hyperinflammatory group and negative values indicating positive enrichment in Hypoinflammatory group. Top 20 significant pathways are shown. AA = amino acid; FA = fatty acid; LC = long chain; MUFA = monounsaturated fatty acid

At both timepoints, the Hyperinflammatory phenotype exhibited significantly reduced levels of lysophospholipids, lysoplasmalogens, plasmalogens, sphingomyelins, phosphatidylcholines, phosphatidylethanolamines, and diacylglycerols compared to Hypoinflammatory phenotype (**Fig. S3**). Conversely, the Hyperinflammatory phenotype maintained high levels of lipid pathways related to the metabolism of steroids; dicarboxylate monohydroxy, long chain monounsaturated, and medium chain fatty acids (FAs); primary and secondary bile acids; and endocannabinoids. Persistent elevations were also observed across amino acid metabolic pathways; TCA cycle intermediates; pentose and aminosugar metabolism; nicotinate/nicotinamide metabolism; and purine/pyrimidine nucleotide metabolism. Three pathways initially elevated in the Hyperinflammatory phenotype at Day 0— glycolysis/gluconeogenesis/pyruvate metabolism, fructose/mannose/galactose metabolism, and methionine/cysteine/SAM/taurine metabolism—were instead significantly reduced compared to the Hypoinflammatory phenotype by Day 2. Notably, lactoyl amino acids and sterols demonstrated differential abundance only at Day 0, while eicosanoids, vitamin C metabolism, and glutathione metabolism were differentially abundant exclusively at Day 2.

In patients surviving through Day 2, individual metabolite trajectories did not differ by 90-day mortality in the full cohort or within phenotypes, except for cinnamoylglycine and a pregnenolone steroid (**Fig. S4**). However, several metabolic classes demonstrated significantly different trajectories based on 90-day mortality (**Fig. S5A**). Tryptophan metabolism, steroid pathways, and gamma-glutamyl amino acids increased over time in non-survivors, who also demonstrated decreasing levels of lactosylceramides, lysoplasmalogens, hexosylceramides, sphingolipids, phospholipids, and ascorbate/aldarate metabolites. Hypoinflammatory non-survivors had increasing progestin steroids (**Fig. S5B**), while Hyperinflammatory non-survivors exhibited decreasing acyl carnitines, plasmalogens, and ascorbate/aldarate metabolites alongside rising pregnenolone steroids (**Fig. S5C**).

### Mitochondrial metabolites are associated with Hyperinflammatory phenotype and mortality

We hypothesized that observed derangements in fatty acid oxidation, lactoyl amino acids, and TCA metabolites stemmed from mitochondrial dysfunction. To test this hypothesis, we curated mitochondria-associated metabolites based on established circulating biomarkers in genetic mitochondrial disorders (*16*). Of 38 detectable mitochondria-associated metabolites in our cohort, 37 (97%) differed significantly between phenotypes (**Fig. 3A**). Since vasopressors can enhance glycolysis and lactate production (*17, 18*), we investigated whether increased mitochondrial metabolic activity in the Hyperinflammatory group merely reflected vasopressor administration. Differential abundance analysis incorporating vasopressor administration (≥ 1 hour infusion in preceding 24 hours) as a covariate revealed that 31 (81%) mitochondrial metabolites remained differentially abundant between phenotypes (**Fig. 3A**), suggesting the distinct mitochondrial signature in the Hyperinflammatory phenotype is independent of vasopressor effects. Further examining metabolic mitochondrial function through plasma redox-coupled (e.g., NADH/NAD^+^) metabolite pairs (*19–23*), we observed both lactate:pyruvate and 3-hydroxybutyrate:acetoacetate ratios were significantly higher in Hyperinflammatory patients (**Fig. 3B**), indicating systemic redox imbalance. Finally, assessing clinical relevance, 26 of 38 mitochondrial metabolites (68%) were associated with 90-day mortality in multivariate logistic regression models (**Fig. 3C**).

**Figure 3.**
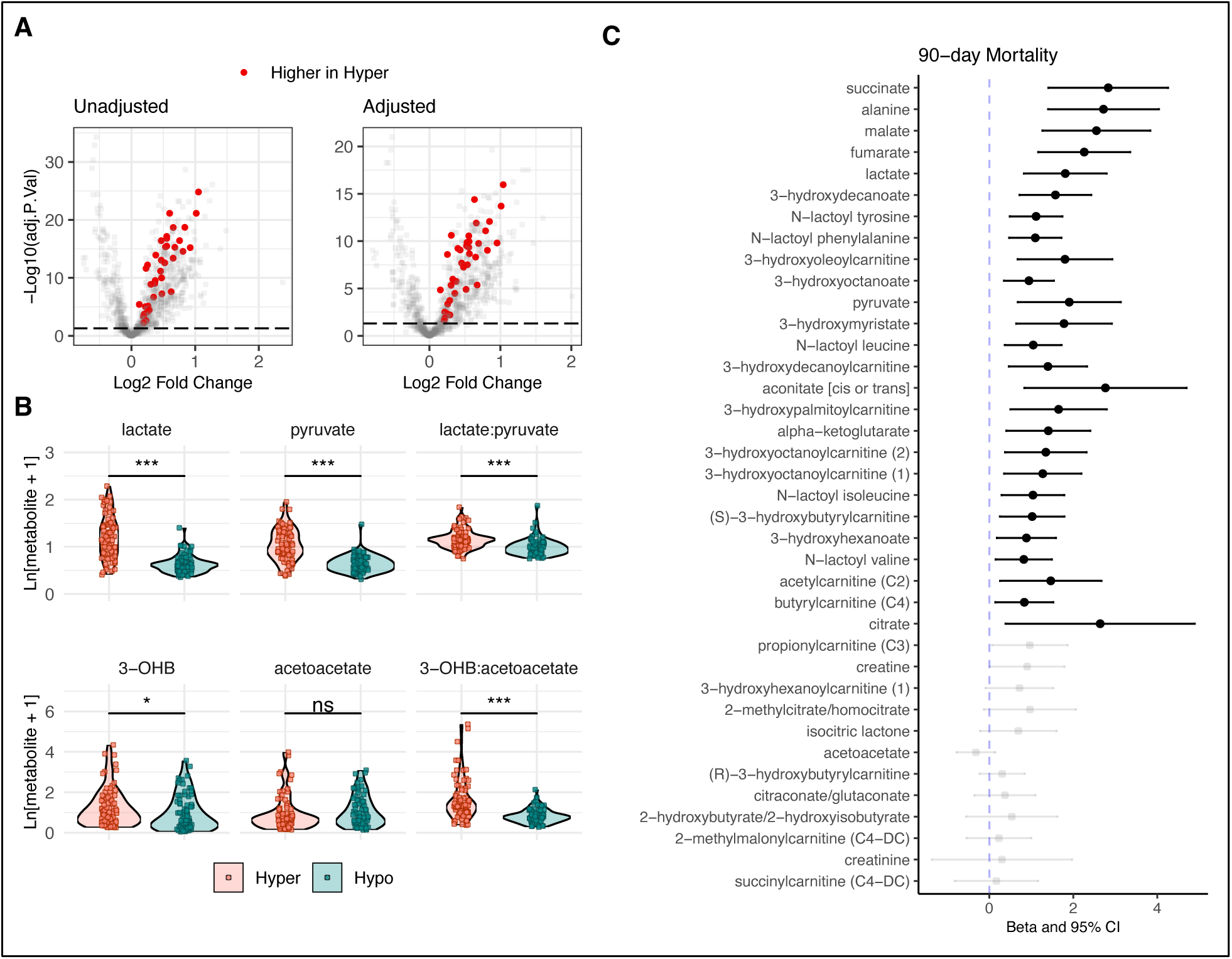
Mitochondrial metabolic derangements across LCA phenotypes. (**A**) Unadjusted: Day 0 volcano plot of metabolites showing statistical significance (limma) vs log2 fold-change comparing hyperinflammatory with hypoinflammatory phenotype. Solid colors represent mitochondrial metabolites. Adjusted: Statistical significance vs log2 fold-change using limma and adjusted for covariates age, sex, BMI, and vasopressor use. (**B**) Peak intensities of redox-coupled mitochondrial metabolites at Day 0. (**C**) Association of mitochondrial metabolites at Day 0 with 90-day mortality in all patients (N = 160). X-axis depicts regression coefficient with 95% confidence intervals derived from logistic regression models using log-transformed peak intensity of each metabolite as the primary predictor and adjusting for covariates age, sex, BMI, and vasopressors. Solid circles represent FDR p-value <0.05. 3-OHB: 3-hydroxybutyrate

### Multi-omics analysis identifies principal factors related to LCA phenotypes

To identify principal sources of biological heterogeneity in the ROSE cohort, we next performed integrated analysis of longitudinal metabolomics and whole blood transcriptomics across all patients (**Fig. 1B**). We selected the top 500 metabolites and 2500 gene transcripts by median absolute deviation in the full cohort for multi-omics analysis (**Fig. 4A**). Applying a MEFISTO (<u>Me</u>thod for the <u>F</u>unctional <u>I</u>ntegration of <u>S</u>patial and <u>T</u>emporal <u>O</u>mics data) model incorporating both data types from both timepoints, we used temporal information as a covariate and configured the model to identify 10 latent factors (*24*). MEFISTO is a dimensionality reduction unsupervised approach for integrating multi-modal data to identify driving sources of variation across data modalities and disentangle sources of variation that change over time from those that are independent of time. Though MEFISTO does not enforce factor orthogonality, Spearman’s correlation analysis revealed no significant inter-factor correlations, confirming each factor captured a distinct source of variability (**Fig. 4B**). The model explained 49.7% of the total variance (*R*^2^) in transcriptomic data and 40.6% in metabolomic data (**Fig. S6**). Factors 1-3 collectively accounted for 58% of explained transcriptomic variance and 72% of explained metabolomic variance (**Fig. 4C, Table S4**). Factor 1 was predominantly driven by transcriptomic data, Factor 2 by metabolomic data, and Factor 3 by both data modalities.

**Figure 4.**
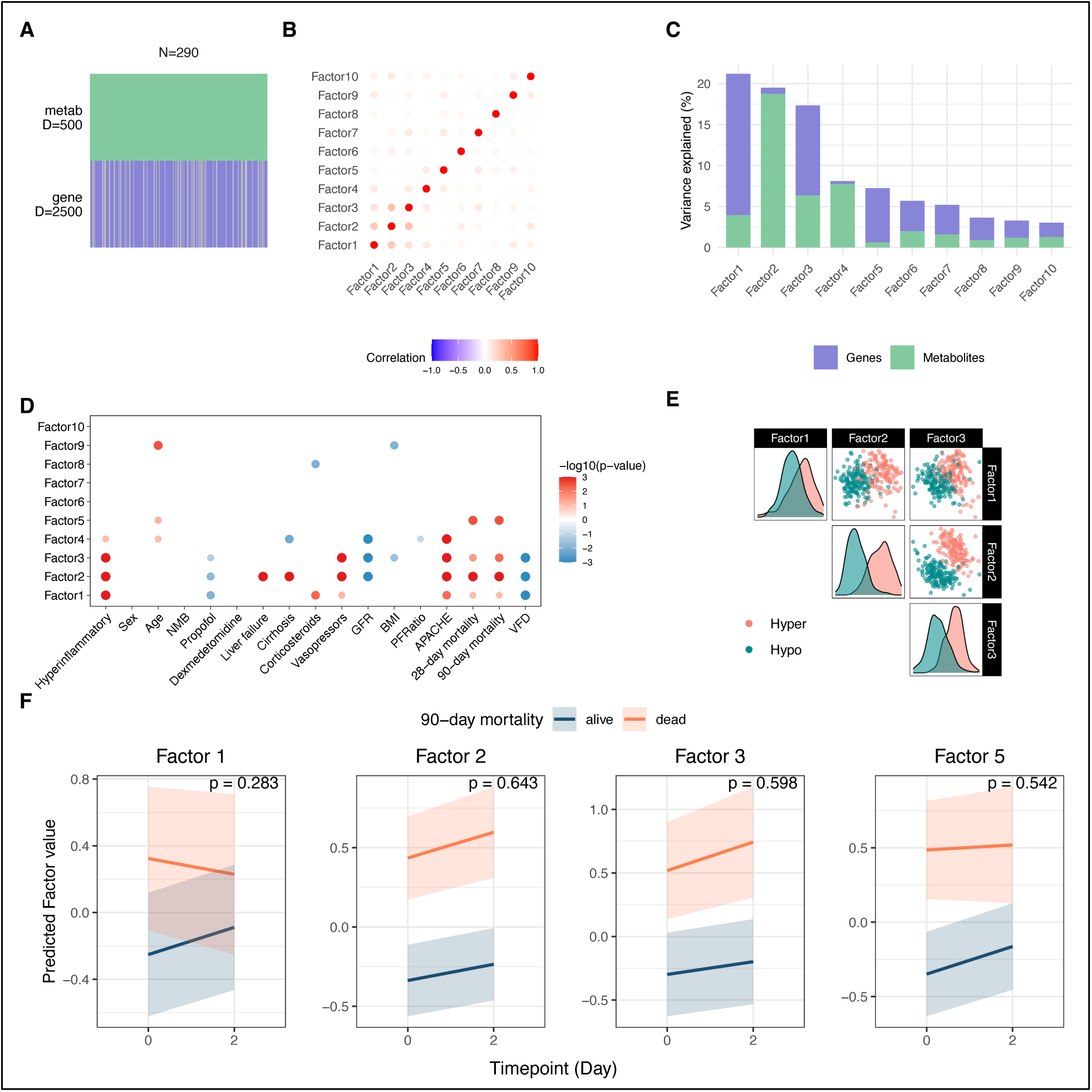
Multi-omics factor analysis (MEFISTO) overview and association with clinical outcomes. (**A**) MEFISTO model overview with 290 samples (from 160 patients across two timepoints), top 500 metabolites and top 2500 gene transcripts by median absolute deviation. (**B**) Spearman’s correlation plot of 10 MEFISTO factors based on factor values. (**C**) Proportion of total variance explained by each factor and each data modality (metabolite vs gene transcript). (**D**) The association of MEFISTO factor values at Day 0 with clinical variables. Size and transparency of the dots represent strength of association as determined via Spearman’s correlation for continuous predictors or linear regression for categorical predictors (FDR <0.05). Color represents directionality of the correlation. (**E**) Paired plots of MEFISTO factor values per patient sample, colored by LCA phenotype designation. (**F**) The slope of change in factor values over time by survival. Only factors associated with mortality at Day 0 are depicted. P-value derived from interaction term of a linear mixed effects regression model with 90-day mortality, timepoint and their interaction as fixed effects and patient as random effect.

We next analyzed associations between each latent factor at Day 0 and key clinical characteristics and outcomes. Factors 1-3 exhibited strong associations with LCA phenotype designation, APACHE III scores, mortality, and ventilator free days, while demonstrating variable associations with GFR, vasopressor use, corticosteroid administration, and propofol infusion (**Fig. 4D**). Notably, Factor 5, lacking association with LCA phenotypes, demonstrated strong independent association with mortality. While Factor 2 substantially separated phenotypes, the combination of Factors 2 and 3 achieved near-complete phenotype discrimination (**Fig. 4E**). These findings indicate that the principal sources of biological heterogeneity identified through our data-driven multi-omic approach strongly aligned with the biological signals captured by LCA phenotype designation.

### Multi-omic factors are related to mortality

Factors 1, 2, 3 and 5 demonstrated strong associations with mortality (**Fig. 4D**). MEFISTO identified all factors as time-independent (time scales = 0), and the rate of change in factor values over time did not differ by 90-day mortality outcome (**Fig. 4F**).

Factor 1, predominantly driven by gene expression, revealed coordinated changes between whole blood transcripts and plasma metabolites, primarily reflecting innate immune activation (**Fig 5A-D**). Gene set enrichment analysis demonstrated significant positive enrichment in neutrophil degranulation and TLR1:TLR2 signaling pathways, glycosaminoglycan (GAG) metabolism, lipid metabolism, and 5-eicosatetraenoic acid (5-ETE) synthesis pathways, with negative enrichment in protein synthesis/trafficking and EIF2AK4-mediated integrated stress response pathways. These transcriptional changes accompanied systemic metabolic alterations characterized by decreased plasma levels of long-chain polyunsaturated fatty acids, lysophospholipids, and plasmalogens, coupled with elevated pregnenolone steroids (precursors for all endogenous steroid production), lactoyl amino acids, glycolytic intermediates, and branched chain amino acid catabolites.

**Figure 5.**
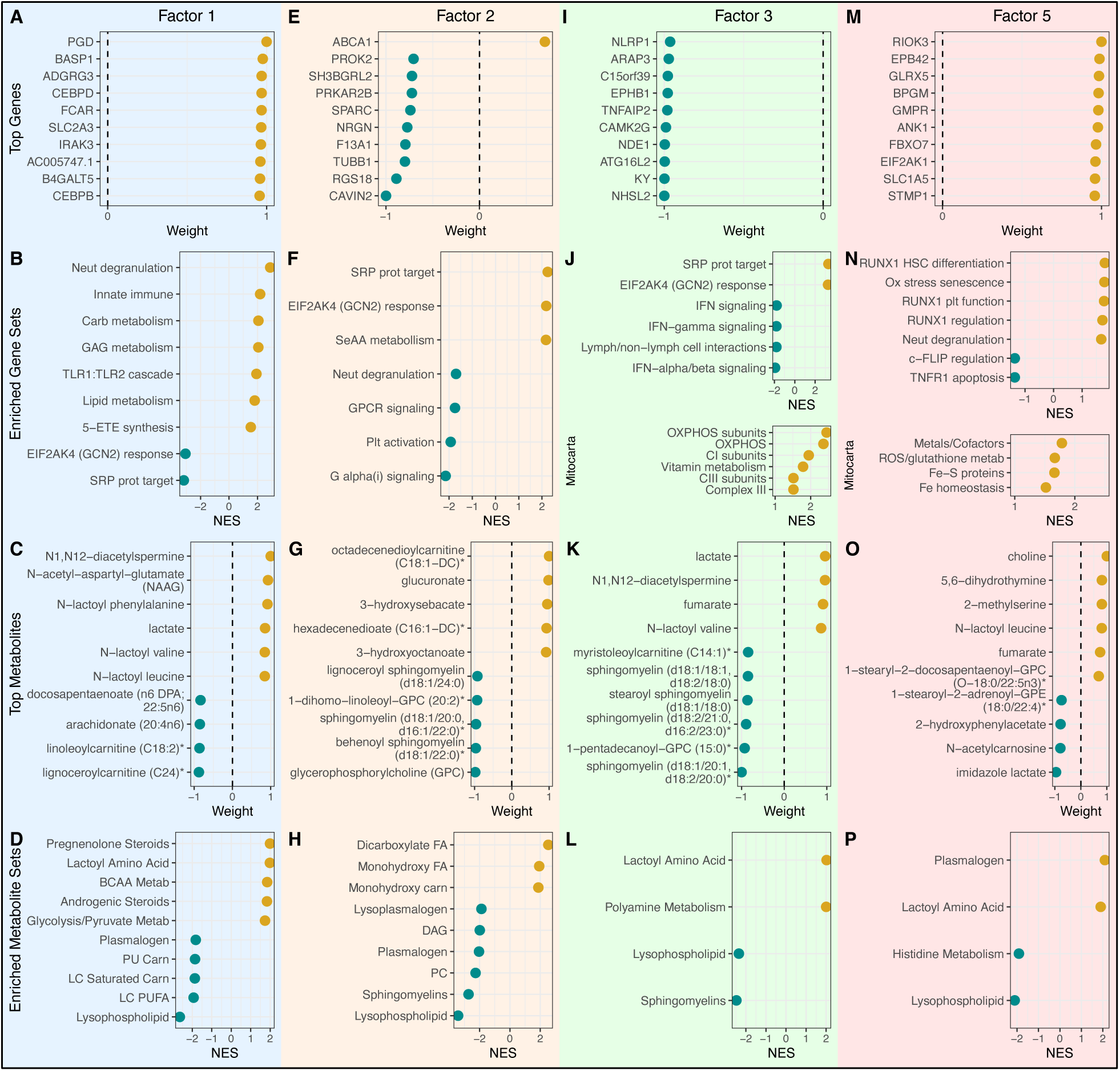
Top features and pathways within mortality-associated MEFISTO Factors. (**A**, **E**, **I**, **M**) Top 10 genes by relative scaled weight in each factor. (**B**, **F**, **J**, **N**) Select top gene set enrichment pathways in each factor using Reactome unless otherwise stated. X-axis depicts normalized enrichment score (NES). (**C**, **G**, **K**, **O**) Top 10 metabolites by relative scaled weight in each factor. (**D**, **H**, **L**, **P**) Top metabolic pathways. X-axis depicts normalized enrichment score (NES).

Factor 2, significantly associated with clinical evidence of hepatic and renal dysfunction (**Fig. 4D**), was primarily metabolite-driven (**Fig. 5E-H**). The plasma metabolome demonstrated accumulation of ω-oxidation products (mono- and di-carboxylated fatty acids) alongside decreased membrane-associated lipids and lipid signaling molecules (**Fig. 5H**). Transcriptional profiling revealed increased expression of ABCA1, the cholesterol efflux pump, as well as positive enrichment of translation machinery and EIF2AK4-mediated amino acid stress response pathways (**Fig. 5F**), with negative enrichment in neutrophil degranulation, platelet activation, and G-protein coupled receptor signaling pathways.

Factor 3, associated with clinical evidence of renal dysfunction (**Fig. 4D**), was characterized by reduced interferon signaling and increased systemic metabolic stress (**Fig. 5I-L**). Transcriptional analysis revealed positive enrichment of basic cellular processes including protein synthesis and RNA processing, while immune signaling pathways were broadly suppressed (**Fig. 5J**). Higher Factor 3 values corresponded with increased expression of mitochondrial oxidative phosphorylation genes, particularly complexes I and III. Notably, both Type I and Type II interferon signaling pathways were downregulated, alongside decreased expression of lymphoid cell interaction genes. These transcriptional changes were accompanied by elevated lactoyl amino acids and polyamines, and reduced sphingomyelins and lysophospholipids.

Factor 5 values were significantly associated with mortality but not LCA phenotype (**Fig. 4D**). Analysis revealed a molecular state characterized by cell proliferation and oxidative stress (**Fig. 4M-P**). Transcriptional profiling demonstrated positive enrichment of DNA replication, cell cycle progression, and RUNX1-mediated hematopoietic differentiation pathways, with negative enrichment of death receptor signaling pathways including TNFR1 and RIPK1 (**Fig. 5N**). This hyperproliferative state featured increased expression of mitochondrial iron homeostasis genes, Fe-S protein metabolism, and ROS management systems, concurrent with activation of oxidative stress-induced senescence pathways.

### Multi-omics analysis reveals mortality-associated signatures within LCA phenotypes

To investigate mechanisms underlying outcome heterogeneity within each ARDS phenotype, we conducted separate multi-omics factor analyses <u>within</u> each phenotype (**Fig. 1C**). Using MEFISTO with identical parameters to our full cohort analysis, we found that, in both phenotypes, transcriptional variation contributed more substantially to within-phenotype heterogeneity than metabolomic variation (**Fig. 6A-B**, **Fig. 7A-B**).

**Figure 6.**
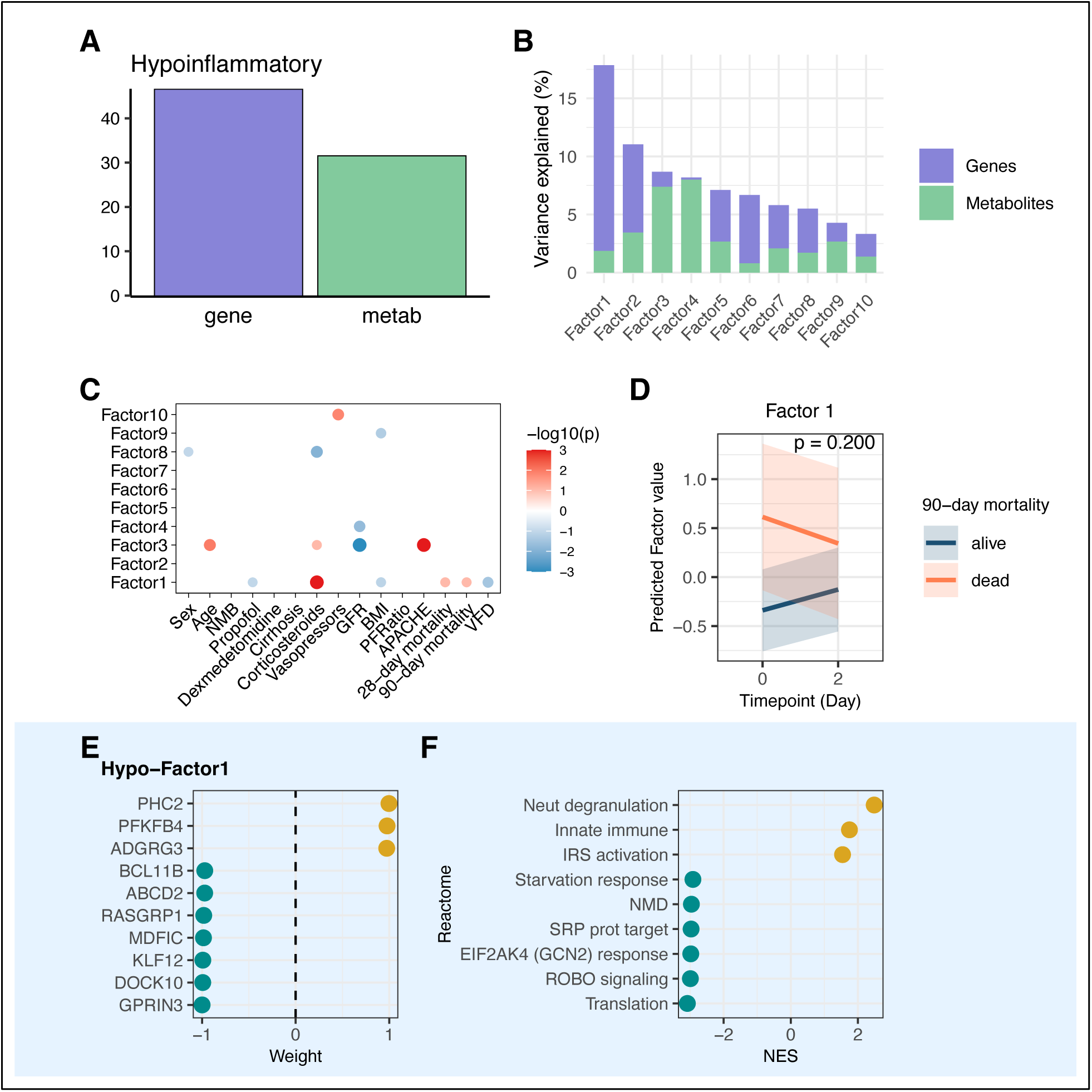
Hypoinflammatory MEFISTO and mortality-associated signature. Analysis of 153 samples from 80 patients at two timepoints. (**A**) Proportion of total variance explained per MEFISTO factor and per data modality (gene, metabolite). (**B**) Proportion of total variance explained per MEFISTO factor. (**C**) Association of MEFISTO factors with clinical variables at Day 0 as determined via Spearman’s correlation for continuous predictors and linear regression for categorical predictors (FDR <0.05). (**D**) The slope of change in factor 1 over time by 90-day mortality. P-value derived from interaction term of a linear mixed effects regression model with 90-day mortality, timepoint and their interaction as fixed effects and patient as random effect. (**E**) Top genes in Factor 1 by relative scaled weight. (**F**) Enriched gene expression pathways in Factor 1. X-axis depicts normalized enrichment score (NES). NMD = nonsense-mediated decay.

**Figure 7.**
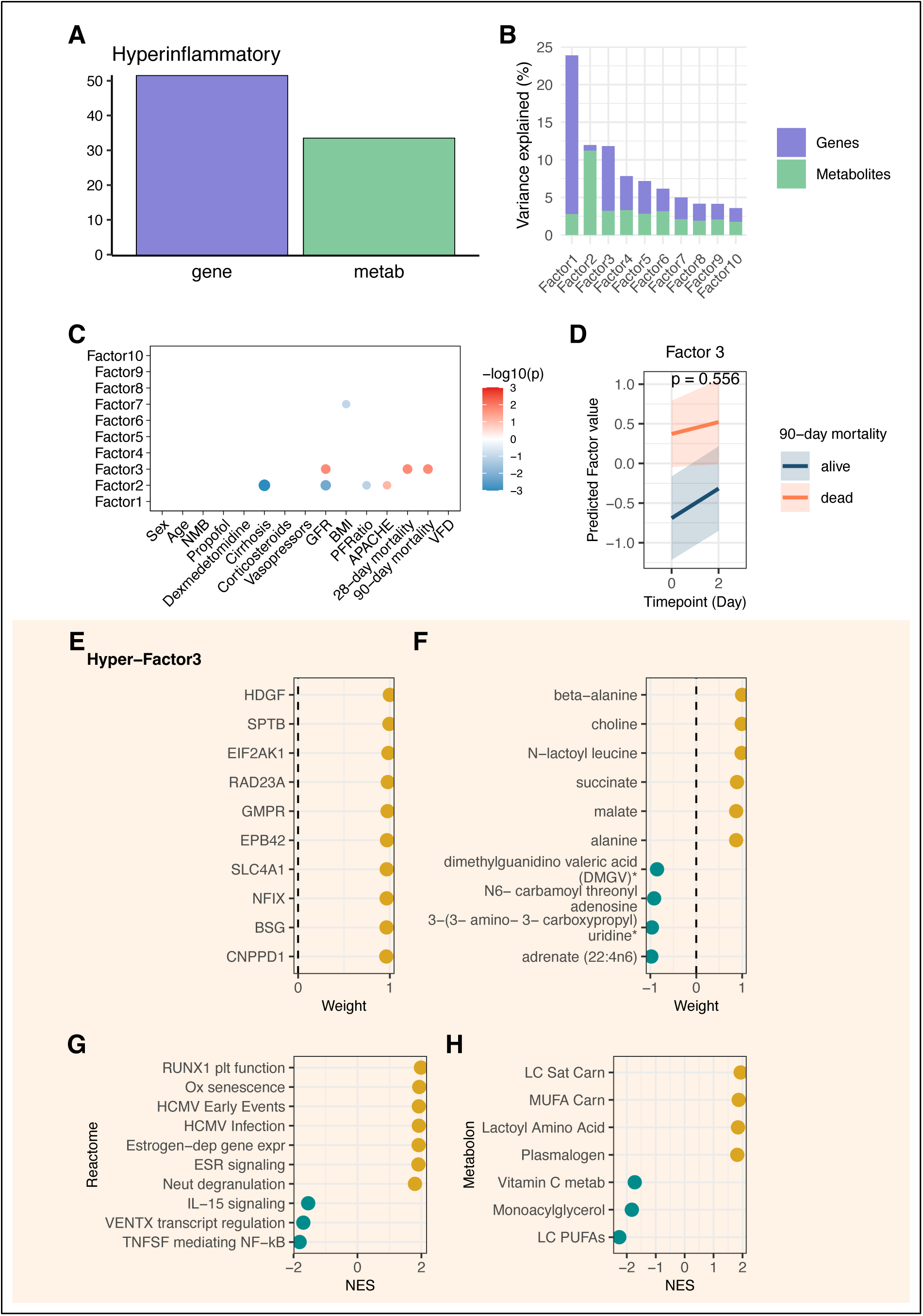
Hyperinflammatory MEFISTO and mortality-associated signature. Analysis of 137 samples from 80 patients at two timepoints. (**A**) Proportion of total variance explained per data modality (gene, metabolite). (**B**) Proportion of total variance explained per MEFISTO factor. (**C**) Association of MEFISTO factors with clinical variables at Day 0 as determined via Spearman’s correlation for continuous predictors and linear regression for categorical predictors (FDR <0.05). (**D**) The slope of change in factor 3 over time by 90-day mortality. P-value derived from interaction term of a linear mixed effects regression model with 90-day mortality, timepoint and their interaction as fixed effects and patient as random effect. (**E**) Top genes in Factor 3 by relative scaled weight. (**F**) Top metabolites in Factor 3 by relative scaled weight. (**G**) Top enriched gene expression pathways by normalized enrichment score (NES) in Factor 3. (**H**) Top enriched metabolic pathways by NES in Factor 3. ESR: estrogen receptor; LC: long chain; metab: metabolism; MUFA: monounsaturated fatty acid; plt: platelet; PUFA: polyunsaturated fatty acid. Sat: saturated.

In the Hypoinflammatory group, Factor 1, primarily characterized by gene expression patterns, was associated with mortality (**Fig. 6B-C**). Factor 1 values demonstrated no differential change over time based on survival status (**Fig. 6D**) but had strong association with moderate-to-high dose corticosteroid treatment in the preceding 24 hours. Gene expression profiling revealed positive enrichment of innate immune response pathways, neutrophil degranulation, and IRS activation with concurrent negative enrichment of starvation response, nonsense mediated decay, and integrated stress response pathways amongst others (**Fig. 6E-F**).

In the Hyperinflammatory group, Factors 1-3 explained most data variance (**Fig. 7B**). While Factor 1 had no association with clinical variables, Factor 3 demonstrated strong association with mortality, with similar temporal trajectories between survivors and non-survivors (**Fig. 7C-D**). Factor 3 was characterized by elevated expression of genes involved RUNX1 mediated platelet function, oxidative-stress induced cellular senescence, HCMV infection, estrogen dependent gene expression and signaling, and neutrophil degranulation, with negative enrichment in IL-15 signaling, transcriptional regulation by VENTX, and TNF receptor superfamily mediating non-canonical NF-kB pathways (**Fig. 7E**, **Fig. 7G**). TCA cycle intermediates and mitochondrial metabolites (malate, succinate, fumarate, lactate) were positively weighted (**Fig. 7F**). Metabolite analysis identified systemic stress markers (lactoyl amino acids), altered lipid metabolism, and reduced vitamin C metabolism (**Fig. 7H**).

### Multi-omic signatures are validated in external cohorts

To assess generalizability of MEFISTO latent factors derived from our cohort with extreme phenotype designations, we examined these associations in EARLI, an ongoing prospective observational cohort study of critically ill adults with sepsis (**Fig. 1D**). A subset of EARLI patients meeting sepsis criteria within two days of enrollment (n = 818) had previously undergone LCA phenotyping (*25*). Metabolomic data were available for 195 patients, whole blood transcriptomics for 196 patients, and both data types for 61 patients (**Fig. S7, Table S5**) (*26, 27*).

To project ROSE MEFISTO factors onto EARLI patients, we selected the top 100 highest-weighted features by absolute scaled weight within each factor of interest, yielding two transcriptomic signatures (Factors 1 and 3), and one metabolomic signature (Factor 2). This approach reduced noise from lower-weighted features and enabled testing in a larger cohort. We calculated Factor 1 and 3 scores for EARLI patients with transcriptomic data (n=196) and Factor 2 values for those with metabolomic data (n=195). All three ROSE MEFISTO factors demonstrated similar LCA phenotype associations in EARLI, with improved phenotype discrimination achieved by combining Factors 2 and 3 (**Fig. 8A-C**). The four mortality-associated ROSE MEFISTO factors were similarly associated with mortality in EARLI (**Fig. 8D**).

**Figure 8.**
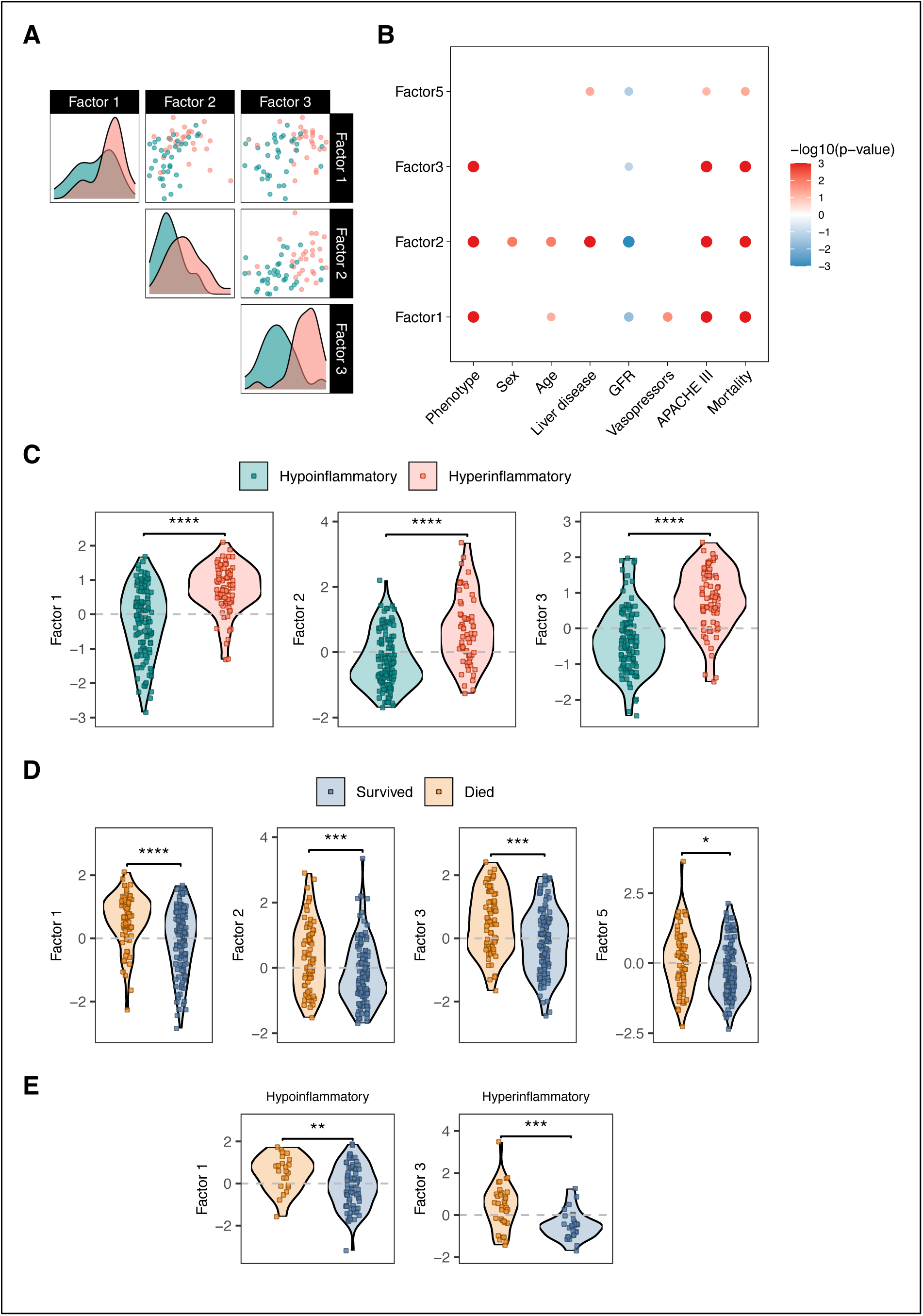
ROSE MEFISTO factor projections in the EARLI observational cohort. (**A**) Scatterplot of paired projected factor values per patient sample, colored by LCA phenotype designation, in patients with both transcriptomic and metabolomic data available (N = 61). (**B**) Association of projected MEFISTO factor values with clinical variables as determined via Spearman’s correlation for continuous predictors and linear regression for categorical predictors. (C) Projected factor values per patient sample comparing Hyperinflammatory to Hypoinflammatory phenotype. P-value determined by Wilcoxon rank sum. N = 189 patients with transcriptomic data (Factors 1 and 3), and N = 183 patients with metabolomic data (Factor 2). (D) Projected factor values per patient sample comparing hospital survivors to non-survivors in those with physician-adjudicated sepsis and transcriptomic data (N = 196 for transcriptomic Factors 1, 3, and 5; N = 195 for metabolomic Factor 2). P-value determined by Wilcoxon rank-sum. (**E**) Projected phenotype-specific MEFISTO factors associated with mortality onto the extreme (p>0.9) Hypoinflammatory (left, N = 101) and extreme Hyperinflammatory (right, N = 61) patients in EARLI. *: p≤0.05, **: p ≤ 0.01, ***: p ≤ 0.001, ****: p ≤0.0001

Using the same approach, we projected ROSE MEFISTO mortality-associated factors derived <u>within</u> each LCA phenotype onto the EARLI participants with high probability (p > 0.9) of belonging to that phenotype. The top 100 features in each phenotype-specific factor yielded one transcriptomic signature per phenotype. Transcriptomic data were available for 101 participants with Hypoinflammatory sepsis and 61 with Hyperinflammatory sepsis (both with phenotype probability >0.9). Both phenotype-specific mortality signatures demonstrated significant mortality associations in the EARLI cohort (**Fig. 8E**).

## DISCUSSION

In this integrated multi-omic analysis of ARDS inflammatory phenotypes, we identified distinct transcriptional and metabolomic signatures that differentiate Hyperinflammatory from Hypoinflammatory phenotypes and are associated with clinical outcomes. Three key insights emerged: First, the Hyperinflammatory phenotype exhibits profound mitochondrial dysfunction and metabolic derangement associated with mortality, persisting independently of vasopressor use, suggesting an intrinsic phenotypic feature. Second, longitudinal multi-omic integration revealed four mortality-associated molecular factors representing distinct pathobiological processes: (1) innate immune activation with enhanced glycolysis, (2) hepatic dysfunction coupled with impaired fatty acid oxidation, (3) suppressed interferon signaling with altered mitochondrial respiration, and (4) maladaptive immune cell proliferation with redox stress. Third, we identified biological signals associated with mortality <u>within</u> each inflammatory phenotype and quantified their relative contribution to overall biological heterogeneity and temporal evolution. These molecular signatures were replicated in an independent cohort of critically ill patients with sepsis, indicating their generalizability. Together, these findings advance our understanding of ARDS and sepsis heterogeneity and identify potential therapeutic targets for phenotype-specific interventions.

Factor 1, accounting for the largest proportion of molecular variation (35% transcriptomic, 10% metabolomic variance), reveals crucial insights into the relationship between inflammation and outcomes in ARDS. This Factor represents an enhanced innate immune response through neutrophil activation and TLR1:TLR2 signaling, coupled with hypermetabolism. The increased expression of genes related to synthesis of inflammatory mediators (5-ETE) combined with reduced plasma PUFA levels suggest active consumption of circulating lipids, likely to support increased energy demands of activated immune cells and generation of lipid mediators. Glycosaminoglycan (GAG) metabolism enrichment suggests tissue remodeling and altered barrier function, while elevated lactoyl amino acids and increased glycolysis suggest widespread mitochondrial metabolic stress and potential Warburg effect, or aerobic glycolysis, as this factor was independent of hypoxia status (PaO2:FiO2) (*16, 28*). Together, these findings suggest a coordinated systemic response where circulating immune cells exhibit inflammatory activation with corresponding metabolic adaptation via increased glycolysis and lipid metabolism. Notably, Factor 1 had the weakest association with mortality, suggesting that interventions solely targeting broad suppression of inflammatory responses may be insufficient to fundamentally reduce mortality related to ARDS and sepsis. Indeed, the stronger signatures of mortality in this cohort were related to Factors 2 and 3, both characterized by attenuated immune responses.

A consistent mortality signal in our analyses was related to renal and hepatic dysfunction coupled with impaired fatty acid *β*-oxidation (Factor 2), strongly associated with the Hyperinflammatory phenotype. Dicarboxylic fatty acids (DCFAs), generated primarily in liver and kidney through *ω*-oxidation, a process excess fatty acids when mitochondrial beta-oxidation is compromised (*29, 30*). Elevated DCFAs, typically detected in urine of patients with mitochondrial fatty acid oxidation disorders, can further impair mitochondrial respiration and ATP synthesis via mitochondrial uncoupling (*31, 32*). The combination of elevated DCFAs and low plasmalogen levels suggests peroxisomal dysfunction, as DCFAs undergo preferential peroxisomal *β*-oxidation, and peroxisomes are essential for plasmalogen biosynthesis (*33, 34*). Peroxisomes also play a crucial role in regulating inflammation by maintaining neutrophil membrane phospholipid composition and viability. Together, this metabolic signature, with its persistent elevation over time in non-survivors, implies liver and kidney dysfunction leading to metabolic derangements that could further exacerbate end-organ dysfunction and contribute to impaired immunity, creating a vicious cycle strongly associated with mortality. Therapeutic interventions targeting lipid homeostasis restoration, such as L-carnitine supplementation, plasmalogen replacement, or simvastatin, could be candidates for study in this patient population (*34–36*).

Factor 3, strongly associated with both the Hyperinflammatory phenotype and mortality, represents broad impairment in interferon signaling (type I and type II) and lymphoid cell interactions, alongside enrichment of integrated stress response pathways, increased cell turnover, and altered mitochondrial respiration. Suppressed type I interferon responses have been documented in peripheral blood of patients with severe COVID-19, in monocytes from bronchoalveolar lavage of patients with COVID-19/metapneumovirus co-infection, and in pediatric patients with severe respiratory syncytial virus infection (*37–39*). Similarly, reduced interferon signaling was observed in the MARS1 transcriptional phenotype of critically ill sepsis patients at highest mortality risk (*40*). Whether this broad interferon program suppression results from pathogen-specific mechanisms or host biological heterogeneity remains unclear. Therapeutic interferon-*γ* has shown promise in sepsis-induced immunosuppression, particularly benefiting patients with decreased monocyte HLA-DR expression and reduced TNF production in response to LPS, and has proven effective in treating fungal sepsis in chronic granulomatous disease and HIV-associated cryptococcal meningitis (*41–43*).

Our current findings validate and deepen our previous work on plasma metabolic profiles in ARDS phenotypes (*14*). While our earlier pilot study identified reduced circulating lipids and elevated glycolytic metabolites in Hyperinflammatory ARDS, our present multi-omic analysis elucidates the mechanistic underpinnings of these derangements. Mitochondrial stress emerged as a central theme across all mortality-associated MEFISTO factors, with lactoyl amino acids— recently established biomarkers of mitochondrial dysfunction in inherited metabolic disorders and predictors of septic shock mortality—significantly elevated in three of the four factors (*16, 28*). Each factor highlighted distinct perturbations in mitochondrial bioenergetics coupled with specific immune signatures: Factor 1 revealed metabolic reprogramming suggestive of the Warburg effect alongside enhanced innate immunity; Factor 2 demonstrated specific deficits in fatty acid *β*-oxidation with impaired immune responses related to liver dysfunction; Factor 3 highlighted increased expression of oxidative phosphorylation and electron transport chain genes coupled with interferon program suppression; and Factor 5 identified mitochondrial redox imbalance with immune cell proliferation and oxidative stress-induced cellular senescence. The metabolic signatures, together with broad depletion of membrane lipids across all factors, offer mechanistic explanations for the reduced circulating lipids previously observed in our work and independent sepsis cohorts (*44, 45*). This molecular dissection of ARDS heterogeneity demonstrates the intricate interplay between mitochondrial bioenergetics and immunophenotype, suggesting combination therapies targeting both metabolic derangements and inflammation may achieve synergistic reductions in ARDS and sepsis mortality. Notably, previous experimental work identified mitochondrial dysfunction in alveolar epithelial type 2 cells that was rescued by mitochondrial transfer from mesenchymal stromal cells, resulting in recovered surfactant secretion and reduced lung injury severity—highlighting the therapeutic potential of interventions restoring mitochondrial function (*46*).

Our phenotype-specific multi-omic analyses reveal that within-phenotype biological heterogeneity had modest associations with mortality. Rather, the primary biological differences driving outcome variation were those that distinguish the inflammatory phenotypes from each other. Nevertheless, examination of mortality-associated signatures within each inflammatory phenotype uncovered distinct mechanistic patterns. In Hypoinflammatory ARDS, the mortality signature strongly correlated with moderate-to-high dose corticosteroid use within the preceding 24 hours and was characterized by enhanced innate immunity coupled with reduced expression of genes related to the integrated stress response and starvation response. Since steroid administration in ROSE was clinician-directed rather than protocol-driven, this association may reflect confounding by indication. Without comprehensive data on steroid dosing and duration, this relationship cannot be interpreted as causal. However, existing evidence suggests patients with Hypoinflammatory-like phenotypes may respond poorly to corticosteroids, as demonstrated in a secondary analysis of the VANISH trial, where the Hypoinflammatory phenotype experienced worse outcomes when randomized to corticosteroids (*47, 48*). These findings suggest that steroid responsiveness may vary significantly among ARDS phenotypes, underscoring the necessity for phenotype-stratified clinical trials to optimize therapeutic approaches.

In Hyperinflammatory ARDS, mortality was associated with increased expression of genes related to platelet activation and neutrophil degranulation, estrogen-dependent signaling, and CMV viral infection coupled with reduced NK/T cell programs suggested by reduced in IL-15 signaling and non-canonical NF-kB2 pathways. Non-survivors exhibited enhanced RUNX1-mediated transcriptional regulation governing hematopoietic stem cell and megakaryocyte differentiation. Notably, RUNX1 knockdown attenuates inflammatory cytokine production in LPS-stimulated macrophages, while its inhibition mitigates proinflammatory mediators and improves survival in septic shock models (*49, 50*). Similarly, endothelial progenitor cells-derived exosomes silencing RUNX1 ameliorated sepsis-induced AKI in experimental models (*51*).

Lastly, metabolomic analyses revealed depleted circulating long-chain polyunsaturated fatty acids, likely from oxidative stress-induced peroxidation and consumption of inflammatory lipid mediators, accompanied by elevated plasmalogens and long chain acyl carnitines indicative of impaired fatty acid beta-oxidation. Collectively, these data suggest mortality in the Hyperinflammatory phenotype results from multifactorial dysregulation spanning innate and adaptive immunity, platelet activation, lipid metabolism, and estrogen signaling pathways.

Our findings offer several clinical implications. The identification of mortality-associated molecular signatures presents opportunities for targeted interventions based on specific biological mechanisms. These signatures remain stable during the initial 48 hours post-ICU admission, providing a potential therapeutic window. Our data indicate multiple contributing pathways to mortality, suggesting combination therapies may yield synergistic benefits, similar to IL-6 inhibitors with dexamethasone in COVID-19 related ARDS (*52*). With emerging precision medicine platform trials in critical care and point-of-care phenotyping tools for inflammatory phenotypes, therapeutics targeting these signatures can be systematically evaluated across phenotypes (*13, 53*).

Our study has several key strengths that enhance the robustness and generalizability of its findings. To our knowledge, our analysis, which used samples from 160 patients in the multicenter ROSE trial, represents the largest multi-omics analysis in ARDS to date. The clinical trial framework ensured standardized care and systematic biospecimen collection, minimizing treatment-related confounding. Our multi-modal approach comprehensive insights into cellular programming and systemic metabolism in ARDS and sepsis. The two-timepoint design established signature stability, critical for therapeutic target identification. Lastly, external validation in a diverse sepsis cohort indicates these molecular states represent generalizable traits of critical illness, aligning with evidence that ARDS inflammatory phenotypes extend to sepsis(*25*) and overlap with other protein and transcriptional subtypes (*40, 48, 54, 55*).

Important limitations include the inability of observational human biospecimen data to establish causality between identified signatures and outcomes. Whole blood transcriptomics precludes attribution of gene expression patterns to specific immune cell populations, while untargeted metabolomics provides limited source information for the observed differences in circulation, which may include liver, kidney, and lung. The absence of comprehensive pathogen data restricts contextualizing these molecular signatures within the broader pathophysiology of ARDS and sepsis. Finally, clinical utility of these molecular signatures requires further investigation in both experimental models and clinical studies.

In conclusion, this comprehensive multi-omic analysis reveals novel insights into the molecular heterogeneity of ARDS and sepsis. Inflammatory phenotypes of ARDS and sepsis reflect distinct biological processes with profound differences in mitochondrial function, immune response, and metabolic regulation. Mortality-associated molecular states suggest complex interplay between phenotype-specific and phenotype-independent pathways affecting patient outcomes. Future studies must determine tissue origins of these circulating signatures, the impact of specific pathogens, and test viable therapeutic targets in experimental models, laying groundwork for interventions that address the molecular complexity of critical illness.

## MATERIALS AND METHODS

### Study design and cohorts

#### Primary Cohort

The ROSE randomized trial of neuromuscular blockade for moderate-to-severe ARDS enrolled 1,006 patients from January 2016 to April 2018 (*15*). Patients were randomized to continuous cisatracurium infusion with deep sedation versus usual care, with the trial stopping early due to futility for the primary outcome of 90-day mortality. LCA of clinical and protein biomarker data was previously performed on all patients with Day 0 biospecimens available, with participants assigned probabilities of membership to Hyper- or Hypo-inflammatory phenotypes (*10*). We randomly selected 80 patients from each phenotype who had a >0.9 probability of phenotype membership. This sample size was determined *a priori* to enable detection of differences between survivors and non-survivors within each phenotype, assuming mortality rates of 40% in Hyperinflammatory and 20% in Hypoinflammatory ARDS based on prior studies (*5, 11, 56–58*). Using the *MetSizeR* package with probabilistic principle components analysis (PPCA) and a fixed FDR of 0.05, this sample size (32 predicted deaths in Hyperinflammatory and 16 in Hypoinflammatory) provided >90% power to detect metabolic differences via untargeted profiling (*26, 59, 60*). Samples were obtained from the NHLBI biorepository, BioLINCC.

#### Validation Cohort

The <u>E</u>arly <u>A</u>cute <u>R</u>enal and <u>L</u>ung <u>I</u>njury (EARLI) study is an ongoing prospective observational cohort of critically ill adults admitted to ICUs at the University of California San Francisco Moffitt-Long Hospital and Zuckerberg San Francisco General Hospital. Patients are eligible upon ICU admission from the emergency room, excluding those with isolated neurological/neurosurgical indications or trauma service admissions. The University of California San Francisco Institutional Review Board approved this study. From this cohort, we analyzed three partially overlapping subgroups (**Fig. S3**): 195 patients with sepsis (2008-2016) who previously underwent metabolic profiling (*26*); 196 participants with hypotension or requiring invasive mechanical ventilation in the emergency room and sepsis (2010-2018) who previously underwent transcriptomic profiling (*27*); and 308 patients from 818 sepsis patients (2008-2019) who underwent LCA of clinical and protein biomarker data (*25*). This subset of 308 patients was selected because they had both LCA phenotype designation and either transcriptomic data, metabolomic data, or both available. Sepsis diagnosis was adjudicated through retrospective physician review of electronic medical records using sepsis-2 criteria, incorporating all available clinical and microbiologic data while blinded to phenotype or biological profiling data (*61*). Patients whose initial sepsis diagnosis occurred >2 days after ICU admission were excluded.

### Biomarker measurements

#### Metabolic profiling

EDTA plasma (150 uL) from Day 0 and Day 2 of ROSE trial enrollment was batch shipped to Metabolon (Durham, NC), precipitated with methanol, and underwent untargeted metabolic profiling using three complementary methods: reverse phase chromatography/ultra performance liquid chromatography tandem mass spectrometry (RP/UPLC-MS/MS) with positive electrospray ionization (ESI), RP/UPLC-MS/MS with negative ESI, and hydrophilic interaction liquid chromatography (HILIC)/UPLC-MS/MS with negative ESI. Metabolon performed peak identification using an in-house library in 2023, as well as quality control and batch-normalization.

In EARLI, 150 uL of citrated plasma underwent identical untargeted profiling methodology, with peaks identified using Metabolon’s in-house library in 2017 (*26*).

#### RNA sequencing

In the ROSE cohort, whole blood samples from Day 0 and Day 2 of trial enrollment were collected in PAXgene tubes, stored at −80°C, and RNA extracted using Qiagen RNEasy kit followed by DNAse treatment as previously described (*10*). In EARLI, whole blood RNA sequencing was performed using a similar methodology (*27*).

### Statistical analysis

Analyses were conducted in R version 4.3.2. Clinical variables and demographics were compared between the phenotypes using Welch’s t-test, Wilcoxon rank-sum test, Chi-squared test, or Fisher’s exact test as appropriate based on variable type, distribution, and expected frequency.

For metabolomic analyses, unknown metabolites and those with >25% missingness in both phenotypes were removed. Following Kokla et al.’s approach to minimize imputation error (*62*), metabolites with >25% missingness in either phenotype were imputed using a uniform distribution ranging from ½ minimum to minimum observed value of the metabolite across all samples. The remaining metabolites were imputed using Random Forest (*missForest*). Metabolic profiles were compared via differential abundance analysis using limma (*MetaboAnalystR* package), adjusting for age, sex, BMI, relevant medications (propofol, dexmedetomidine, corticosteroids), comorbid liver disease, and GFR (*63*). For Day 2 analyses, randomization arm was added as a covariate, as samples were obtained after the administration of trial agents. Metabolite enrichment analysis was performed using ChemRICH (*64*), a chemical similarity-based statistical enrichment approach that overcomes limitations of traditional pathway analysis. By grouping metabolites based on chemical ontologies and structural similarity, ChemRICH generates study-specific, non-overlapping metabolite sets with self-contained enrichment statistics independent of background database size. For our analysis, differentially abundant metabolites at each timepoint (Day 0 or Day 2) with their identifiers (SMILES, InChIKeys, PubChem IDs) and Metabolon class annotations. After resolving duplicate entries and completing missing PubChem IDs through database searches, the dataset was processed through the ChemRICH web interface.

For each metabolite, we constructed linear mixed-effects models to analyze changes in metabolite values over time based on 90-day mortality outcome. The primary model included fixed effects for time, mortality, treatment arm, age, sex, and BMI, with a random intercept for each subject. We tested the significance of the time-by-mortality interaction by comparing this model to a null model without the interaction term using likelihood ratio tests. The coefficient of the time-by-mortality interaction represents the differential trajectory of metabolite levels between survivors and non-survivors from Day 0 to Day 2, with positive values indicating greater increases (or smaller decreases) in non-survivors. P-values from model comparisons (FDR <0.05) were used to assess statistical significance of these differential trajectories. For analysis of metabolic class trajectories over time, we annotated differentially expressed metabolites with pathway information from Metabolon’s database. Fold changes were calculated by exponentiating the model coefficients and adding 1, representing the relative change in metabolite levels between survivors and non-survivors. We performed enrichment analysis using ChemRICH as described above. Significantly enriched pathways (FDR<0.05) were classified as increased or decreased based on the proportion (>0.5) of increased metabolites within each pathway.

For multi-omics analyses in the full study cohort, we implemented a rigorous filtration pipeline to select only the most abundant and variable analytes, thereby avoiding imputation which can introduce artifacts in integrated multi-omics analyses. As such, unknown metabolites and xenobiotics were removed. Metabolites with >10% missingness were removed. Remaining metabolites underwent log transformation, quantile normalization, and selection of the top 500 by median absolute deviation (MAD), followed by z-scaling. Transcriptomic data underwent variance stabilizing transformation, with the top 2500 genes selected by MAD and subsequently z-scaled. For each patient, Day 0 and Day 2 metabolite and gene expression data were entered into a MEFISTO model (*mofa2* package) (*24*). MEFISTO is an unsupervised multi-modal temporally informed dimensionality reduction tool to identify predominant patterns of variation in omics data. MEFISTO extends conventional matrix factorization by incorporating a functional view on latent factors based on Gaussian processes, allowing for modeling of temporal relationships in the data. Our implementation treated the entire patient cohort as a single group while declaring time as a covariate, facilitating joint decomposition of multi-omics data matrices into latent factors (Z) with corresponding feature weights (W), with temporal structure modeled through a squared exponential covariance function. This framework allowed for identification of both smooth (time-dependent) and non-smooth (time-independent) variation patterns, providing insights into temporal dynamics of molecular responses in ARDS patients while accounting for cohort level heterogeneity. After model fitting, the resulting factor values (Z) were extracted to quantify the strength of each identified molecular co-variation pattern for each patient at each timepoint, allowing us to characterize the temporal dynamics of metabolomic and transcriptomic responses in ARDS patients.

We selected 10 latent factors for initial analysis. The total variance (*R*^2^) explained for each data modality and per factor was calculated to determine the primary sources of dataset heterogeneity. To determine the association of MEFISTO factors with clinical variables, we performed linear regression for categorical predictors (with factor value as the outcome) and Spearman’s correlation for continuous predictors using Day 0 factor values. Gene set enrichment analysis was performed on latent MEFISTO factors using Reactome and MitoCarta 3.0 gene sets, while a metabolite set was generated using Metabolon’s annotated library (*65, 66*). To test for interaction between MEFISTO factors and time regarding mortality, we implemented linear mixed effects regression models with 90-daymortality, timepoint, and their interaction as fixed effects, including a random intercept for each patient to account for within-subject correlation in measurements over time.

For multi-omics analyses within each LCA phenotype, the same data processing pipeline was applied with MAD-based selection of metabolites and gene transcripts performed within each phenotype.

For validation studies in EARLI, the same pipeline was applied to prepare metabolite and transcriptomic data. The relative weights of the top 100 features within each MEFISTO latent factor that were present in EARLI were used to calculate factor values for each EARLI patient. Specifically, factor values were calculated as the weighted sum of normalized feature measurements, using weights derived from our original MEFISTO model. Associations between factor values per patient and clinical outcomes were tested using Wilcoxon rank sum tests.

All analyses were adjusted for multiple comparisons using the Benjamini-Hochberg false discovery rate (FDR) with significance set at FDR <0.05.

## Data Availability

Transcriptomic data for the ROSE trial participants is in submission to BioDataCatalyst. Metabolomic data will be released to the NIH Metabolomics Workbench database upon acceptance of this manuscript to a peer reviewed journal. Clinical data and biospecimens from the ROSE trial are available through the NHLBI BioLINCC repository (https://biolincc.nhlbi.nih.gov/studies/) to qualified researchers upon request and completion of appropriate data use agreements.

## Acknowledgments

We gratefully acknowledge the ROSE trial investigators, the Prevention and Early Treatment of Acute Lung Injury (PETAL) Network, and the National Heart, Lung, and Blood Institute (NHLBI) Biologic Specimen and Data Repository Information Coordinating Center (BioLINCC) for providing the data and biospecimens that made this research possible. We also gratefully acknowledge the participants and participants’ families in both the ROSE and EARLI studies.

