## Supplementary material for "Longitudinal multi-omic signatures of ARDS and sepsis inflammatory phenotypes identify key pathways associated with mortality": Fig. S1

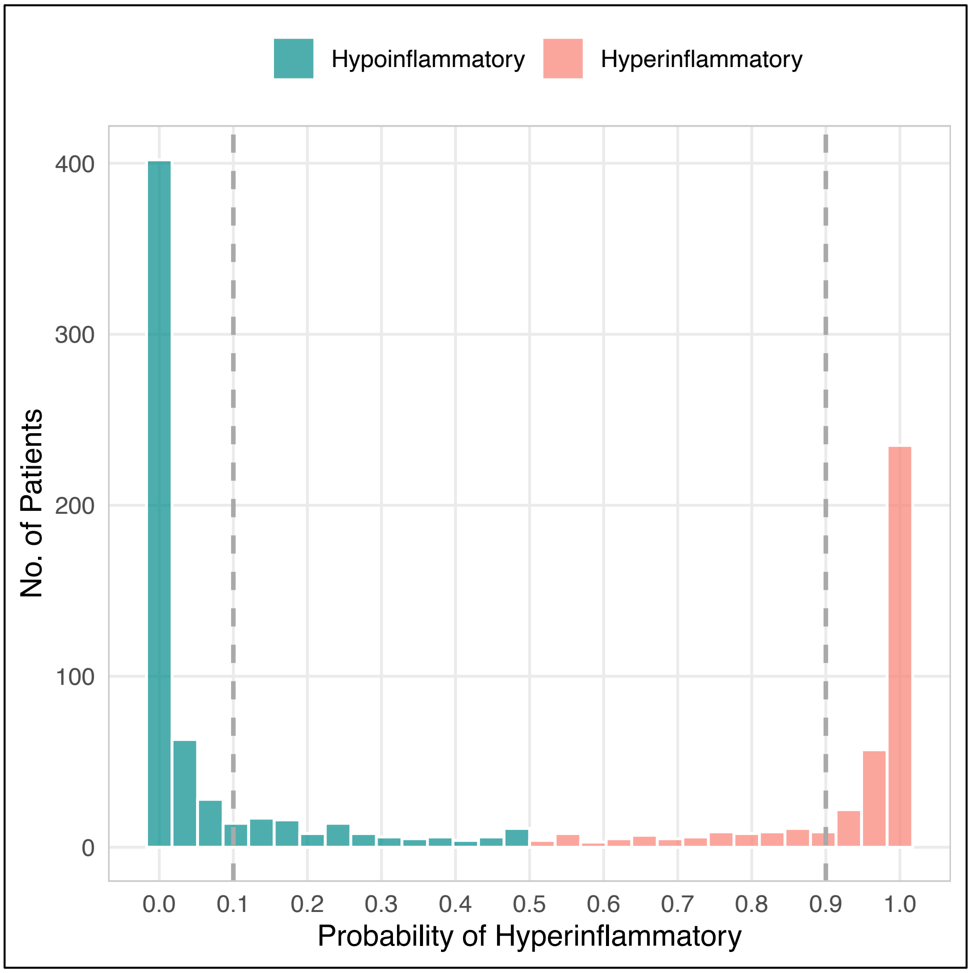


**Figure S1. Distribution of phenotype probabilities from latent class analysis of 1006 patients in the ROSE trial.** Vertical dashed lines indicate the 0.1 and 0.9 thresholds, highlighting regions of high classification certainty into the Hyperinflammatory or Hypoinflammatory phenotypes.
