## Supplementary material for "Longitudinal multi-omic signatures of ARDS and sepsis inflammatory phenotypes identify key pathways associated with mortality": Fig. S2

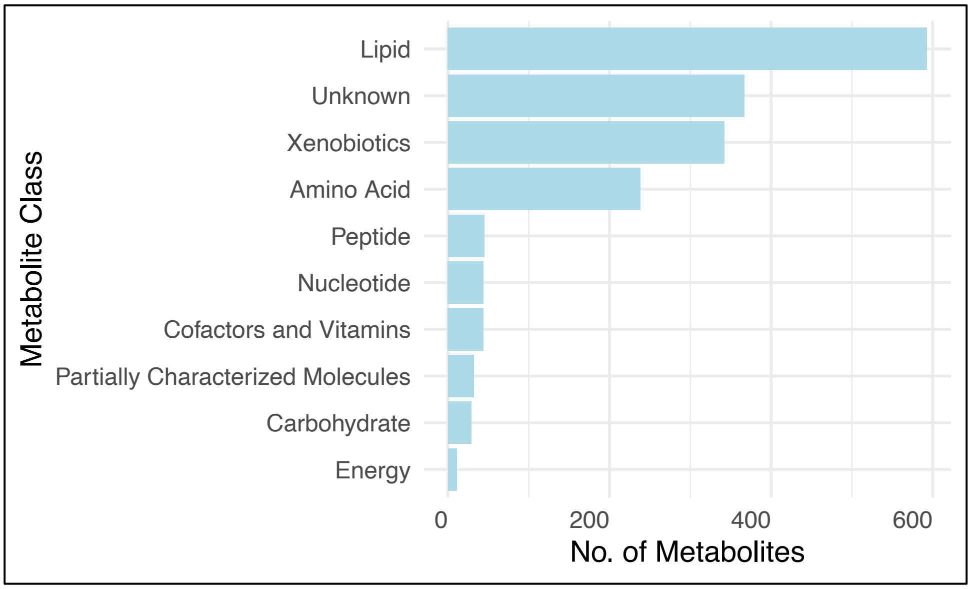


**Figure S2. Results of untargeted metabolic profiling of EDTA plasma from 160 ROSE trial participants.**
