## Supplementary material for "Longitudinal multi-omic signatures of ARDS and sepsis inflammatory phenotypes identify key pathways associated with mortality": Fig. S3

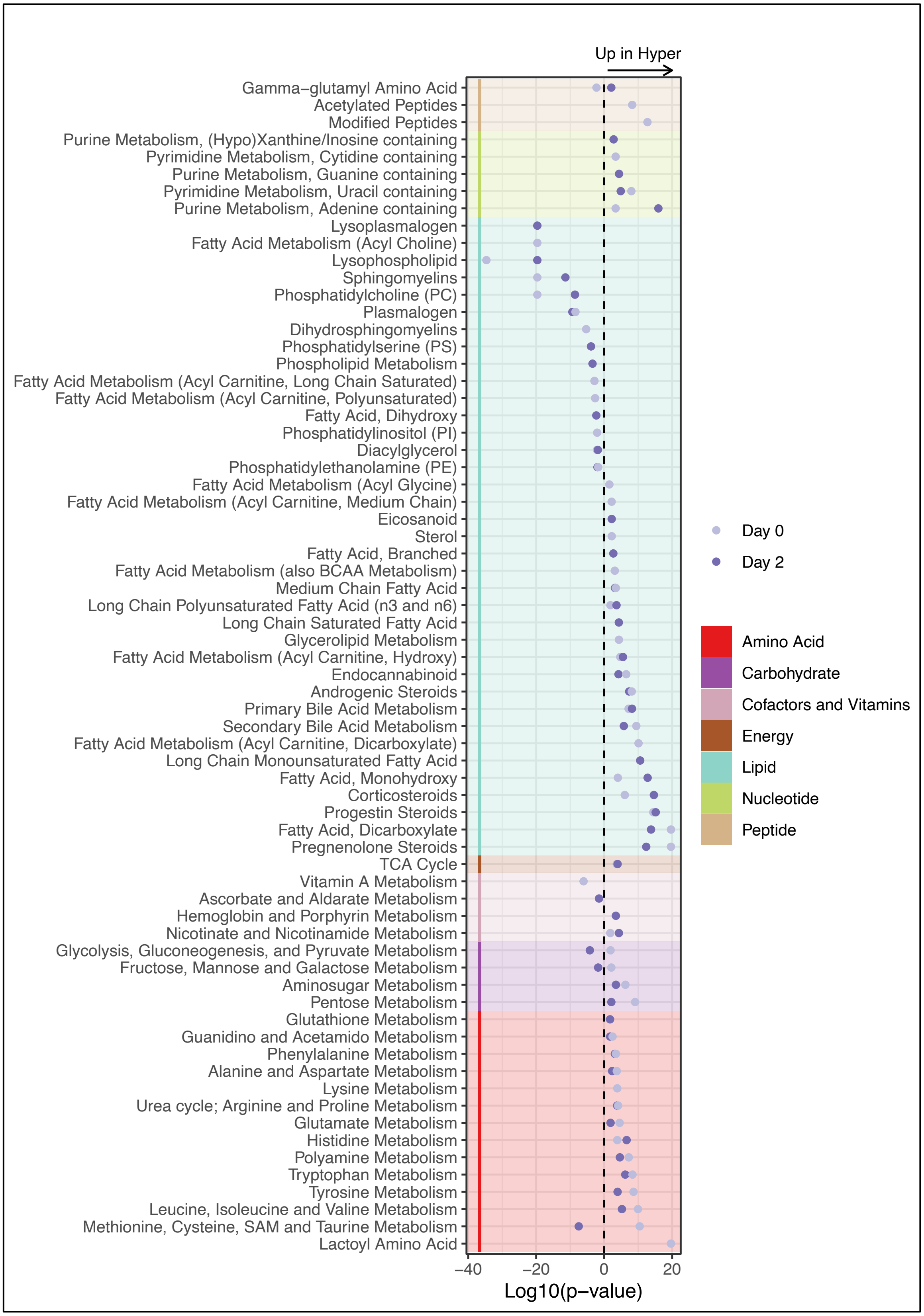


**Figure S3. Enrichment analysis of differentially abundant metabolites by LCA phenotype in the ROSE cohort.** Only significant pathways are displayed (FDR <0.05). Xenobiotics and partially characterized molecules are not depicted.
