## Supplementary material for "Longitudinal multi-omic signatures of ARDS and sepsis inflammatory phenotypes identify key pathways associated with mortality": Fig. S4

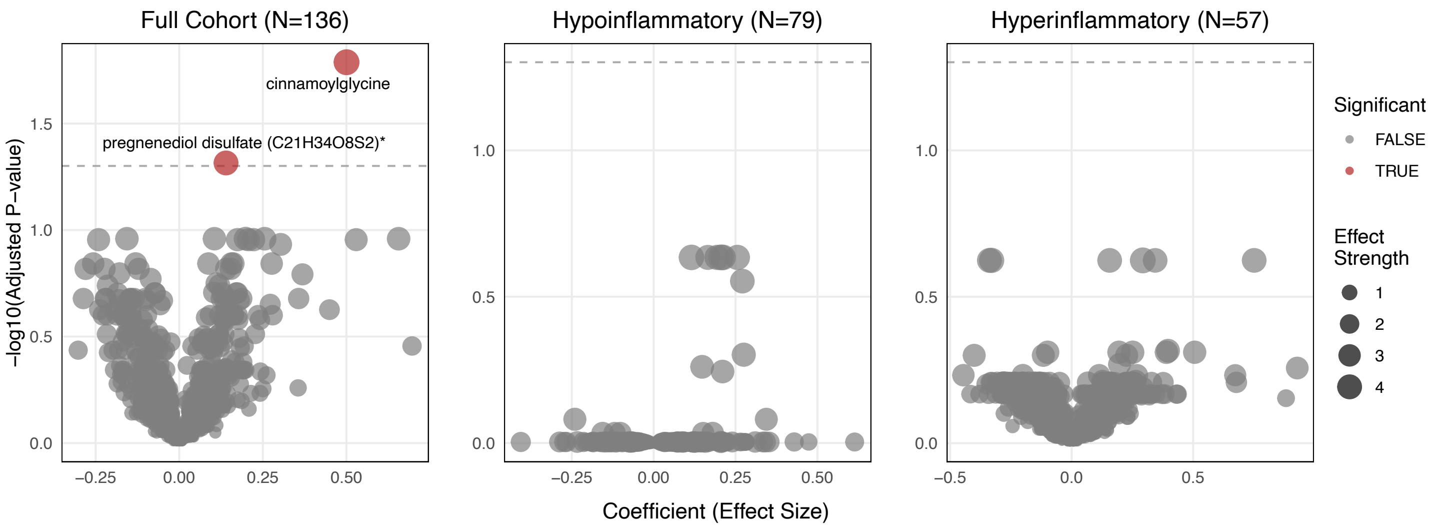


**Figure S4. Metabolite trajectories in 90-day non-survivors vs survivors.** Analysis restricted to the subset who survived through Day 2 post-randomization. Plots display coefficients from linear mixed effects models testing the time-by-mortality interaction, controlling for randomization arm, age, sex, and BMI, and subject-level random effects. Points are sized by effect strength (|coefficient|/standard error), with significance determined by likelihood ratio tests against a model without the interaction term (FDR <0.05). Cinnamoylglycine and pregnenediol disulfate demonstrate the only significant divergent trajectories between survival groups, with stronger positive temporal trends in non-survivors.
