## Supplementary material for "Longitudinal multi-omic signatures of ARDS and sepsis inflammatory phenotypes identify key pathways associated with mortality": Fig. S5

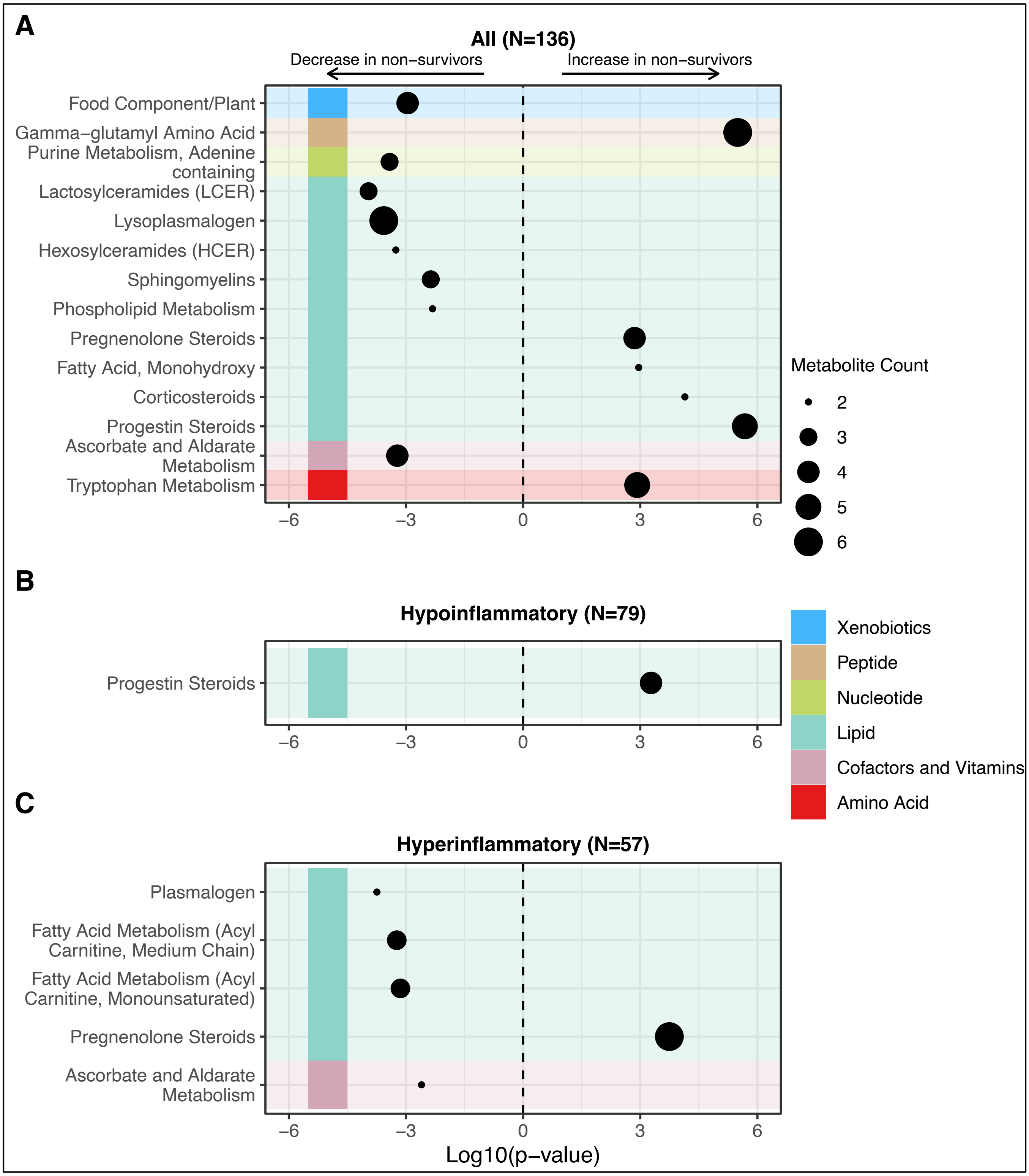


**Figure S5. Differential metabolite pathway enrichment in 90-day non-survivors versus survivors.** Analysis restricted to the subset who survived through Day 2 post-randomization. Linear mixed effects models were used to determine differential rates of change over time controlling for randomization arm, age, sex, and BMI, and subject-level random effects. Model coefficients were converted to fold changes for enrichment analysis. Only metabolic classes with FDR adjusted $p < 0.05$ are depicted. X-axis represents signed statistical significance (Log10[p-value]), with negative values indicating decreased abundance over time in non-survivors relative to survivors, and positive values indicating increased abundance. The size of each dot represents the number of metabolites contributing to each class.
