## Supplementary material for "Longitudinal multi-omic signatures of ARDS and sepsis inflammatory phenotypes identify key pathways associated with mortality": Fig. S6

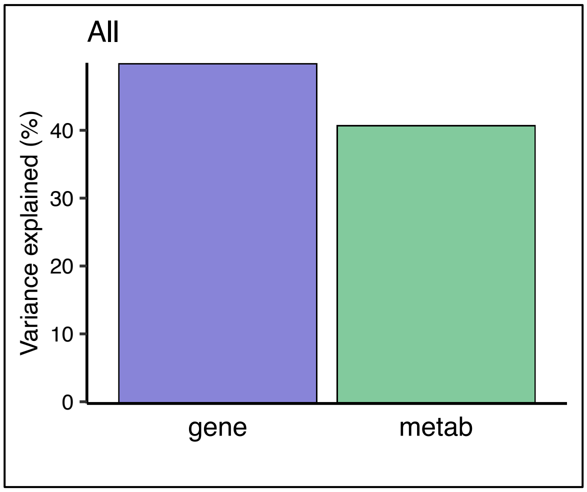


**Figure S6**. **Variance explained per data modality in the ROSE MEFISTO model.** Plots depict proportion of total variance explained per data modality after applying MEFISTO to the full study cohort, n=160**.**
