## Supplementary material for "Longitudinal multi-omic signatures of ARDS and sepsis inflammatory phenotypes identify key pathways associated with mortality": Fig. S7

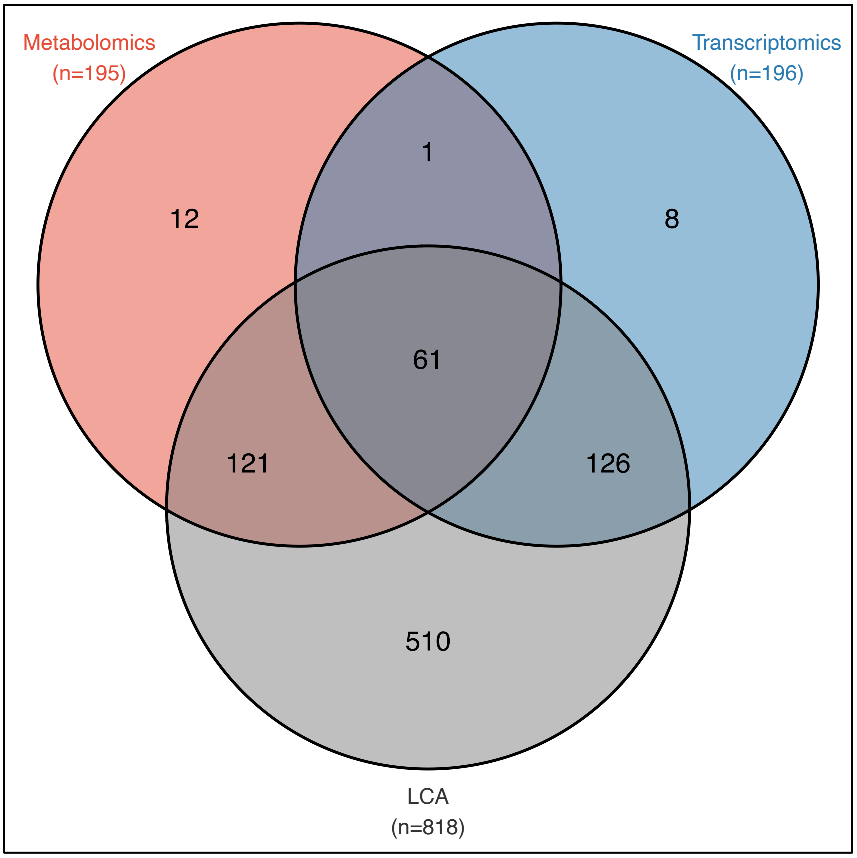


**Figure S7. EARLI validation cohort of patients with sepsis.** Data collection included three overlapping groups: (1) transcriptomic data from study participants with hypotension or receiving invasive mechanical ventilation in the emergency room and sepsis (n=196); (2) protein biomarker data at enrollment available and used for latent class analysis (LCA; n=818); and (3) untargeted metabolic profiling from a randomly selected subset of sepsis patients with and without ARDS (n=195). Only patients with a diagnosis of sepsis within 2 days of enrollment in EARLI were included.
