## Supplementary material for "Longitudinal multi-omic signatures of ARDS and sepsis inflammatory phenotypes identify key pathways associated with mortality": Table S1

**Table S1: Characteristics of patients in the ROSE discovery cohort**

| | Hypoinflammatory (N=80) | Hyperinflammatory (N=80) | Total (N=160) | P-value* |
|---|---|---|---|---|
| **Age**, median (IQR) | 58.5 (46 to 66) | 58.5 (48 to 69) | 58.5 (47 to 68) | 0.38 |
| **Male** | 56 (70%) | 46 (58%) | 102 (64%) | 0.14 |
| **BMI**, median (IQR) | 33 (27 to 38) | 28 (23 to 35) | 30 (25 to 37) | 0.01 |
| **Race** | | | | 0.17 |
| African American | 10 (12%) | 4 (5%) | 14 (9%) | |
| White | 61 (76%) | 64 (80%) | 125 (78%) | |
| Other | 3 (4%) | 1 (1%) | 4 (2%) | |
| Missing | 6 (7%) | 11 (14%) | 17 (11%) | |
| **Randomization arm** | | | | 0.75 |
| Neuromuscular blockade | 42 (52%) | 39 (49%) | 81 (51%) | |
| Usual care | 38 (47%) | 41 (51%) | 79 (49%) | |
| **APACHEIII** | 84 $\pm$ 24 | 132 $\pm$ 31 | 110 $\pm$ 37 | <0.0001 |
| Missing | 30 (37%) | 22 (27%) | 52 (32%) | |
| **PaO2:FiO2[†],** median (IQR) | | | | |
| Lowest in 24 hours | 95 (76 to 126) | 81 (67 to 105) | 86 (71 to 115) | 0.01 |
| At enrollment | 119 (92 to 140) | 116 (93 to 138) | 117 (92 to 140) | 0.70 |
| **GFR**, median (IQR) | 78 (50 to 99) | 26 (13 to 43) | 48 (21 to 86) | <0.0001 |
| **Comorbidities** | | | | |
| Acute kidney injury | 12 (15%) | 42 (52%) | 54 (34%) | <0.0001 |
| Hepatic failure with coma or encephalopathy | 0 | 7 (9%) | 7 (4%) | 0.006 |
| Cirrhosis | 4 (5%) | 15 (19%) | 19 (12%) | 0.007 |
| Chronic dialysis | 2 (2%) | 5 (6%) | 7 (4%) | 0.28 |
| Diabetes | 20 (25%) | 25 (31%) | 45 (28%) | 0.45 |
| Hypertension | 48 (60%) | 42 (52%) | 90 (56%) | 0.47 |
| Congestive heart failure | 6 (7%) | 5 (6%) | 11 (7%) | 1 |
| Prior stroke | 7 (9%) | 5 (6%) | 12 (7%) | 0.78 |
| Chronic pulmonary disease | 26 (32%) | 8 (10%) | 34 (21%) | |
| Immunosuppression within the past 6 months | 12 (15%) | 12 (15%) | 24 (15%) | 1 |
| Leukemia | 0 | 6 (7%) | 6 (4%) | 0.01 |
| AIDS | 2 (2%) | 2 (2%) | 4 (2%) | 1 |
| **Primary cause of ARDS** | | | | <0.0001 |
| Pneumonia | 49 (61%) | 36 (45%) | 85 (53%) | |
| Sepsis[‡] | 0 | 25 (31%) | 25 (16%) | |
| Aspiration | 20 (25%) | 12 (15%) | 32 (20%) | |
| Multiple transfusion | 3 (4%) | 2 (2%) | 5 (3%) | |
| Trauma | 5 (6%) | 1 (1%) | 6 (4%) | |
| Other | 3 (4%) | 4 (5%) | 7 (4%) | |
| **Medications** | | | | |
| Vasopressors | 17 (21%) | 69 (86%) | 86 (54%) | <0.0001 |
| Propofol | 68 (85%) | 48 (60%) | 116 (72%) | 0.002 |
| Dexmedetomidine | 14 (17%) | 11 (14%) | 25 (16%) | 0.77 |
| Corticosteroids[§] | 19 (24%) | 19 (24%) | 38 (24%) | 1 |
| **Ventilator free days**, | 20 (0 to 24) | 0 | 0 (0 to 21) | <0.0001 |

| | Hypoinflammatory (N=80) | Hyperinflammatory (N=80) | Total (N=160) | P-value* |
|---|---|---|---|---|
| median (IQR) | | | | |
| **ICU free days,** median (IQR) | 17.5 (0 to 23) | 0 (0 to 8) | 3.5 (0 to 19) | <0.0001 |
| **Hospital free days**, median (IQR) | 12 (0 to 18) | 0 (0 to 0) | 0 (0 to 15) | <0.0001 |
| **28-day mortality** | 19 (24%) | 45 (56%) | 64 (40%) | <0.0001 |
| **90-day mortality** | 19 (24%) | 49 (61%) | 68 (42%) | <0.0001 |

Numbers are presented as n (%) or mean ± SD unless otherwise stated.
* Determined via Welch's t-test for normally distributed continuous variables, Wilcoxon rank-sum for non-normally distributed continuous variables, and Chi-squared test or Fisher's exact test for categorical variables.
† Lowest PaO2:FiO2 in the 24 hours preceding randomization, and PaO2:FiO2 on day of study enrollment.
‡ If both pneumonia and sepsis were causes of lung injury, pneumonia was reported as primary.
§ Intravenous or enteral corticosteroids (≥ 20 mg methylprednisolone equivalents)
