## Supplementary material for "Longitudinal multi-omic signatures of ARDS and sepsis inflammatory phenotypes identify key pathways associated with mortality": Table S2

**Table S2. Proportion of differentially abundant metabolites between inflammatory phenotypes based on limma results at Day 0.**

| SUPER_PATHWAY | total_count* | significant_count† | proportion_significant |
|---|---|---|---|
| Energy | 11 | 8 | 0.73 |
| Carbohydrate | 27 | 18 | 0.67 |
| Nucleotide | 40 | 24 | 0.60 |
| Lipid | 507 | 295 | 0.58 |
| Amino Acid | 210 | 119 | 0.57 |
| Peptide | 31 | 17 | 0.55 |
| Partially Characterized Molecules | 22 | 11 | 0.50 |
| Cofactors and Vitamins | 35 | 15 | 0.43 |
| Xenobiotics | 99 | 34 | 0.34 |

* Total count reflects the total number of metabolites within a given metabolic pathway entered into the analysis (after removal of metabolites with low variance across the cohort). † Significant count denotes the number of metabolites with FDR <0.05 as identified by limma adjusted for covariates age, sex, BMI, medications, comorbid liver disease, and GFR comparing Hyperinflammatory to Hypoinflammatory phenotypes
