## Supplementary material for "Longitudinal multi-omic signatures of ARDS and sepsis inflammatory phenotypes identify key pathways associated with mortality": Table S3

**Table S3. Proportion of differentially abundant metabolites between inflammatory phenotypes based on limma results at Day 2.**

| SUPER_PATHWAY | total_count* | significant_count† | proportion_significant |
|---|---|---|---|
| Energy | 11 | 7 | 0.64 |
| Partially Characterized Molecules | 22 | 14 | 0.64 |
| Carbohydrate | 27 | 17 | 0.63 |
| Lipid | 507 | 293 | 0.58 |
| Cofactors and Vitamins | 35 | 19 | 0.54 |
| Nucleotide | 40 | 21 | 0.52 |
| Amino Acid | 210 | 83 | 0.39 |
| Xenobiotics | 99 | 35 | 0.35 |
| Peptide | 31 | 7 | 0.23 |

* Total count reflects the total number of metabolites within a given metabolic pathway entered into the analysis (after removal of metabolites with low variance across the cohort). † Significant count denotes the number of metabolites with FDR <0.05 as identified by limma adjusted for covariates age, sex, BMI, medications, comorbid liver disease, and GFR comparing Hyperinflammatory to Hypoinflammatory phenotypes
