## Supplementary material for "Longitudinal multi-omic signatures of ARDS and sepsis inflammatory phenotypes identify key pathways associated with mortality": Table S4

**Table S4. MEFISTO model variance explained for each data type and by each factor.**

| Factor | Gene_Variance | Gene_Percent_of_Total | Metab_Variance | Metab_Percent_of_Total |
|---|---|---|---|---|
| Factor1 | 17.23 | 34.68 | 3.97 | 9.78 |
| Factor2 | 0.74 | 1.49 | 18.76 | 46.23 |
| Factor3 | 10.99 | 22.12 | 6.35 | 15.65 |
| Factor4 | 0.36 | 0.72 | 7.74 | 19.09 |
| Factor5 | 6.62 | 13.32 | 0.62 | 1.53 |
| Factor6 | 3.70 | 7.44 | 2.00 | 4.93 |
| Factor7 | 3.62 | 7.29 | 1.58 | 3.89 |
| Factor8 | 2.73 | 5.49 | 0.91 | 2.23 |
| Factor9 | 2.10 | 4.24 | 1.18 | 2.92 |
| Factor10 | 1.75 | 3.52 | 1.28 | 3.15 |
