## Supplementary material for "Longitudinal multi-omic signatures of ARDS and sepsis inflammatory phenotypes identify key pathways associated with mortality": Table S5

**Table S5. Characteristics of participants with biological data in the EARLI validation cohort**

| | Transcriptomic Cohort (N=196) | Metabolomic Cohort (N=195) | Overlapping Cohort (N=62) |
|---|---|---|---|
| **Age** | 65 ± 15 | 67 ± 14 | 69 ± 11 |
| **Race** | | | |
| White | 75 (38%) | 104 (53%) | 27 (43%) |
| African American | 34 (17%) | 24 (12%) | 11 (18%) |
| Asian | 60 (31%) | 55 (28%) | 20 (32%) |
| Native American | 1 (0.5%) | 0 | 0 |
| Other | 23 (12%) | 12 (6%) | 4 (6%) |
| Unknown | 3 (1%) | 0 | 0 |
| **Male sex** | 118 (60%) | 107 (55%) | 37 (60%) |
| **BMI** | 24 (21 to 29) | 24 (20 to 28) | 24 (21 to 28) |
| **Patient Category** | | | |
| Medicine | 175 (89%) | 159 (81%) | 53 (85%) |
| General Surgery | 3 (1%) | 7 (4%) | 2 (3%) |
| Cardiothoracic Surgery | 1 (0.5%) | 1 (0.5%) | 1 (2%) |
| Vascular Surgery | 1 (0.5%) | 0 | 0 |
| Transplant Surgery | 0 | 1 (0.5%) | 0 |
| Neurology | 0 | 1 (0.5%) | 0 |
| Cardiology | 8 (4%) | 8 (4%) | 2 (3%) |
| Other | 8 (4%) | 18 (9%) | 4 (6%) |
| **Comorbidities** | | | |
| Congestive heart failure | 30 (15%) | 29 (15%) | 9 (14%) |
| Cardiovascular disease | 18 (9%) | 21 (11%) | 6 (10%) |
| Hypertension | 88 (45%) | 127 (65%) | 36 (58%) |
| Chronic lung disease | 63 (32%) | 61 (31%) | 22 (35%) |
| Chronic liver failure | 5 (3%) | 3 (1%) | 2 (3%) |
| Cirrhosis | 20 (10%) | 12 (6%) | 5 (8%) |
| Diabetes | 44 (22%) | 55 (28%) | 1 (26%) |
| Malignancy | 35 (18%) | 65 (33%) | 15 (24%) |
| HIV/AIDS | 13 (7%) | 14 (7%) | 8 (13%) |
| Solid organ transplant | 5 (3%) | 9 (5%) | 3 (5%) |
| Chronic immunosuppression | 12 (6%) | 23 (12%) | 5 (8%) |
| Other immunocompromised | 5 (3%) | 18 (9%) | 1 (2%) |
| ESRD | 11 (6%) | 12 (6%) | 5 (8%) |
| Dialysis | 14 (7%) | 9 (5%) | 4 (6%) |
| CKD | 21 (11%) | 36 (18%) | 8 (13%) |
| **Sepsis type** | | | |
| Pulmonary and non-pulmonary | 12 (6%) | 18 (9%) | 3 (5%) |

| | Transcriptomic Cohort (N=196) | Metabolomic Cohort (N=195) | Overlapping Cohort (N=62) |
|---|---|---|---|
| Non-pulmonary | 70 (36%) | 53 (27%) | 16 (26%) |
| Pulmonary | 108 (55%) | 113 (58%) | 41 (66%) |
| Unclear source | 6 (3%) | 11 (6%) | 2 (3%) |
| **ARDS, Berlin** | 82 (42%) | 88 (45%) | 37 (60%) |
| **ARDS Severity*** | | | |
| Mild | 16 (8%) | 6 (3%) | 3 (5%) |
| Moderate | 39 (20%) | 47 (24%) | 21 (34%) |
| Severe | 27 (14%) | 35 (18%) | 13 (21%) |
| **Acute kidney injury** | 98 (50%) | 108 (55%) | 39 (63%) |
| **GFR** | 56 (26 to 89) | 50 (26 to 81) | 56 (31 to 76) |
| **On vasopressors** | 81 (41%) | 52 (27%) | 24 (39%) |
| **APACHE III** | 115 ± 38 | 100 ± 40 | 121 ± 35 |
| **SAPS II** | 63 ± 20 | 56 ± 22 | 67 ± 19 |
| **LCA Phenotype Assigned** | 187 (95%) | 182 (93%) | 61 (98%) |
| Hyperinflammatory | 76 (39%) | 61 (31%) | 27 (43%) |
| Hypoinflammatory | 111 (57%) | 121 (62%) | 34 (55%) |
| **Outcome** | | | |
| Died | 71 (36%) | 72 (37%) | 31 (50%) |
| Survived | 125 (64%) | 123 (63%) | 31 (50%) |

Numbers are presented as n (%) for categorical variables and mean ± SD for continuous variables.
* Severity defined using the PaO2:FiO2 or the SpO2:FiO2 if the former was missing.
